# “I felt shamed and blamed”: An exploration of the parental lived experience of School Distress

**DOI:** 10.1101/2023.02.16.23286034

**Authors:** Sinéad L. Mullally, Sophie E. Connolly

## Abstract

School Distress refers to a child or young person’s (CYP) difficulty attending school due to the extreme emotional distress they experience before/during/after school. Limited research exists on the impact of School Distress on the parents/carers supporting these CYP. Using a case–control, concurrent embedded mixed-method design, we explored this lived experience. 947 parents of CYP with School Distress completed a bespoke online questionnaire, alongside two control parent groups (n=149, n=25) and one professional group (n=19).

Findings revealed a devastating impact on the mental health of parents, with parents displaying significantly heightened daily anxiety and significantly lower mood during, but not before, their children’s school attendance difficulties. In addition, parents with children experiencing School Distress reported significantly more negative emotion states and significantly fewer positive emotion states. Parents also reported overwhelmingly negative treatment from professionals, including being disbelieved or blamed for their children’s difficulties, threatened with fines and court action, and disempowered by the actions of professionals surrounding their child. Significant, deleterious impacts were also evident across all aspects of their lives, including their careers, finances, and other children. Perhaps unsurprisingly, half of these parents reported developing a new mental health condition since their child’s difficulties began, with the experience itself rated as the second most threatening potential life event, superseded only by the death of a first-degree relative (including a child or spouse). On the other hand, professionals working with CYP with School Distress did not experience these deleterious mental health or wider life consequences. Despite understanding how threatening the experience is for parents, they were often quick to blame parents for their children’s difficulties. Professionals, like parents, expressed frustration with the lack of help available for these CYP and their families.

This study highlights a bleak, adversarial, and lonely picture for parents of CYP struggling to attend school. More specifically, the findings depict a system rife with parental blame; a system that appears to isolate parents through hostile, threatening, and punitive actions. A wider lack of societal understanding of the experience of School Distress further compounds this dearth of support for parents, placing parental mental health in further peril.

## 1. Introduction

### 1.1. School Distress in UK School Children

In 2022, England’s Children’s Commissioner’s attendance audit estimated that, in the 2021 autumn term, 1.7 million pupils in England missed over 10% of school sessions, and 124,000 pupils missed over 50% (1). Updated Department for Education figures show 150,000 pupils (i.e., 1 in every 50 pupils) missed over 50% of school in 2022-23; a figure which is 150% higher than in 2018-19. School absences in Scotland are even higher, with March 2024 figures revealing that 41% of secondary school and 32% of primary school pupils missed over 50% of school in 2022-23 (2). Similarly, in the US, a review of state policy and practice in the school year 2021-22 concluded that chronic absenteeism has blossomed into a full-scale crisis (3). Of additional concern are the findings of the Centre for Social Justice’s termly analysis of official data relating to school absences from English schools, which reported that in the 2023 Autumn term, 1.97% of the school population (i.e., 142,487 pupils) were absent from school more often than they were present (i.e., severely absent); the highest number during an Autumn term on record and 137% higher than before the Covid-19 pandemic (4).

Whilst the underpinnings of these persistent school absences are likely multifaceted, the largest academic study of school attendance difficulties in UK school children to date found that in 94.3% of the cases surveyed, school attendance problems were underpinned by significant emotional distress - with often harrowing accounts of this distress provided by parents (5). This study also reported that, in most of these cases, the children’s School Distress began within their formative years (average age of onset = 7.9 years of age), whilst the average duration was 4 years. Moreover, Connolly et al. (5) also found that children and young people (CYP) struggling to attend school were significantly more likely to have complex neurodivergent profiles than their peers without school attendance difficulties (prevalence = 92.1%; average number of neurodivergences identified = 3.62); with autism (83.4%), multi-modal sensory processing difficulties (56.9%) and ADHD (53.3%) notably prevalent (5).

Comparable findings had previously been reported elsewhere in smaller samples e.g., (6, 7), with a recent survey of parents living in Hackney, London (n=55) conducted by Hackney Independent Parent-Carers (8) closely replicating these findings, with 83% of CYP in this cohort diagnosed autistic and 71% of these autistic CYP having more than one diagnosis. This is an important replication as the cohort here, although relatively small, closely matched the consensus data from the area with respect to ethnicity (57% White British/Other respondents) and household income (with 47% of respondents in receipt of free school meals), whereas Connolly et al.’s (5) was skewed in favour of White British families living in areas with relatively low rates of socio-economic deprivation.

In addition to complex neurodivergent profiles, anxiety is prevalent in these cohorts. For instance, 46% of CYP in Linehan (8) had a diagnosis of anxiety whilst (5) found that 92.5% of the CYP with School Distress demonstrated clinically significant anxiety symptomology. Higher anxiety was also found to correlate with a longer School Distress duration, lower school attendance in the current and previous academic years, and how parents rated the impact that school attendance had on their child’s mental health (with higher anxiety associated with a more severe, negative impact) (5). Elevated demand avoidance (9) was also pervasive (5). Hence, the combined evidence (i.e., the early age of onset, persistent duration, complex neurodivergent profiles, and levels of distress and significant anxiety symptomology in the CYP) suggests that CYP with School Distress are some of society’s most vulnerable CYP, and that they, and their families, are facing significant and persistent daily challenges in the pursuit of education.

### 1.2. Parental Lived Experience of School Distress

In a typical year, English school students are expected to attend school on ∼190 days and, under Section 444 Education Act 1996, their parent(s) have a legal obligation to ensure their attendance, with failure of parents to ensure regular school attendance punishable by a fine of up to £2,500 and a prison sentence of up to 3 months (10). Within the context of School Distress, this inevitably places a significant emotional burden on these parents. Notable, however, there is a concerning dearth of empirical research investigating the parental experience of School Distress. This is likely due to the longstanding discourse in the literature around the topic of School “Refusal”, which posits that persistent school absences are typically due to neglectful, deficient, or failing parenting [e.g., (11, 12); see also (13, 14)].

Such narratives persist, with a recent study of 201 teachers in England indicating that teachers, when asked to describe the causal underpinnings of School Distress, attributed a high level of importance to home (and peer) related factors and least importance to school-related environmental factors (15). This appears to be equally true even in the case of neurodivergence. For instance, using a mixed-method study to explore the school experiences of demand avoidant autistic children via parental report (n=211), Truman and colleagues reported that parents often felt misunderstood by professionals and blamed for their child’s school attendance difficulties (16), up to and including being treated “‘like a criminal and a liar by the school and the education system’” and prosecuted (“‘School number 4 decided to prosecute me instead of helping us’”) (pp.68). Moreover, a survey by Autistic UK (17), that included completed responses from 25 autistic individuals and 224 parent/carers of autistic individuals, again described a high instance of parents being blamed by professionals with reasons such as ‘non-compliance’, ‘overprotective parenting’ and ‘poor parenting’ emerging. They also reported that their respondents’ understanding of the contributing underpinnings of School Distress, such as sensory processing differences, anxiety, and trauma, were often unacknowledged/unrecognised by professionals.

Relatedly, in a thematic analysis of email-based interviews with 40 parents of CYP with school attendance problems who were seeking support for their children, Bodycote (2022) formulated the concept of the ‘Parents Journeys’ (13). This described an overview of common parental experiences, whereby the tension between parental understanding of School Distress and the understanding of school staff and other professionals was front and centre. This tension often led to school staff dismissing parental reports of their children’s difficulties within the school environment, and a tendency for school staff to interpret these difficulties as stemming from deficient parenting abilities and/or problems in the home. Similarly, 62% of parents in Linehan’s (8) Hackney cohort reported feeling judged by school staff, 60% felt ignored, and 59% reported feeling blamed, whilst only 32% reported feeling listened to (8).

The catastrophic consequences of this narrative on parents was amplified by the findings of a survey conducted by Not Fine In School (NFIS) in 2020 (18), which found that 63% of the 714 parents who took part had been blamed for their child’s difficulties, 38% had been reported to social services (with 50 children being put on a Child In Need Plan and 12 children being put on a Child Protection Plan), and 23% (153 respondents) had been accused of Fabricated or Induced Illness (FII) – a form of child abuse whereby a parent or carer exaggerates or deliberately causes symptoms of illness in their child (19) (although in <2% of these cases were parents found guilty, with another 3 cases awaiting a verdict). In addition, parents in this survey frequently reported having been threatened with fines due to schools recording their child’s absence as unauthorised (38%), and a small number were prosecuted for their child’s non-attendance. Again, the children themselves were also often blamed, with 69% of parents reporting that their child was criticised. These testimonies are consistent with the “*pervasive view* [amongst scholars] *that the responsibility of school refusal lies with the individual students and their families*” (14) (pp. 41).

Evidence of the negative impact of school attendance problems on parents has also been highlighted recently by the Local Government and Social Care Ombudsman (LGSCO) (20). Specifically, within one of the case studies presented, the LGSCO described the significant anxiety and distress experienced by parents of a child with school attendance difficulties, who were faced with threats of prosecution when they asked for help. In addition to the above, missed time from work, legal and financial difficulties (21), endangered careers, increased conflict between parents, and parental separations (22) have all been documented in the literature as potential consequences of School Distress for parents. This latter study also highlighted the impact of School Distress on the whole family unit, due to the necessary reorganisation of daily life.

### 1.3. Current Study

Despite the above, the weight of the clinical and academic evidence has focused on parental deficiencies as drivers of school attendance difficulties, with limited research in the psychological literature exploring the impact that having a child who experiences School Distress has on parents. This research sought to address this. By comparing the lived experiences of parents of CYP who have experienced School Distress with both parents of CYP who attend school without distress and parents of CYP who have never attended a school setting, we aimed to qualitatively explore the parental lived experience of School Distress and to quantitively assess the impact on their lives. The questions posed can be considered under five related categories: (1) the direct impact of parenting a child or young person experiencing School Distress on the parent themselves; (2) the interactions that parents have with others, including the professionals/services surrounding the child and family, and how these impact parental lived experiences and mental health; (3) action(s) taken against parents to enforce school attendance and the impact of these actions; (4) key causal factors underpinning School Distress; and 5, key sources of support available to participants supporting CYP with School Distress.

We also recruited a separate group of educational professionals and other professionals who support CYP with School Distress to enable a comparison of the parental experiences and understanding of School Distress with that of professionals in each of the above five categories.

## 2. Method

### 2.1. Participants

#### Parents

Participants were required to live in the United Kingdom and be parents/carers of school aged CYP. 1,121 participants were recruited in total, consisting of 738 parents of children currently experiencing School Distress (Current SD), 209 parents of children who have previously experienced School Distress (Past SD), 149 parents of children who have never experienced School Distress (No SD), and 25 parents of children who have never attended a school setting (Lifelong EHE). 97.03% were mothers. The average completion rate of the questionnaire was 77.35%, with 62.5% of participants completing 100%. A detailed description of the methods and the CYP’s profiles is available elsewhere (5) (see also Table 1).

**Table 1.** Demographic characteristics of the sample. SD =School Distress. EHE = Elective Home Education. ND =Neurodivergence. ADHD = Attention Deficit Hyperactivity Disorder.

| Characteristic | Current SD | Past SD | No SD | Lifelong EHE | Professionals |
| --- | --- | --- | --- | --- | --- |
| *n* | 738 | 209 | 149 | 25 | 19 |
| IMD Decile (StDev) | 6.16 (2.8) | 5.51 (3.1) | 6.17 (2.9) | 5.50 (2.6) | n/a |
| Ethnic Group (%) |  |  |  |  |  |
| White | 458 (93.5%) | 113 (95.8%) | 106 (90.6%) | 16 (94.1%) | 15 (100%) |
| Mixed/Multiple Ethnic Groups | 24 (4.9%) | 4 (3.4%) | 9 (7.7%) | 0 | 0 |
| Asian/Asian British | 3 (0.6%) | 1 (0.8%) | 1 (0.9%) | 1 (5.9%) | 0 |
| Black/African/Caribbean/Black British | 2 (0.4%) | 0 | 1 (0.9%) | 0 | 0 |
| Other Ethnic Group | 3 (0.6%) | 0 | 0 | 0 | 0 |
| Main Language (%) |  |  |  |  |  |
| English | 484 (99.2%) | 117 (100%) | 117 (99.2%) | 17 (100%) | 15 (100%) |
| Other | 4 (0.8%) | 0 | 1 (0.8%) | 0 | 0 |
| Country of Residence (%) |  |  |  |  |  |
| England | 644 (88.0%) | 183 (89.7%) | 129 (86.6%) | 24 (96.0%) | 14 (93.3%) |
| Scotland | 68 (9.3%) | 13 (6.4%) | 13 (8.7%) | 0 | - |
| Wales | 14 (1.9%) | 5 (2.5%) | 2 (1.3%) | 1 (4.0%) | - |
| Northern Ireland | 6 (0.8%) | 3 (1.5%) | 5 (3.4%) | 0 | 1 (6.67%) |
| % Respondents who are Parents | 100% | 100% | 100% | 100% | 80% |
| Respondent who are... |  |  |  |  |  |
| Mothers (%) | 707 (96.6%) | 200 (98.0%) | 145 (97.3%) | 25 (100%) | 10 (66.7%) |
| Fathers (%) | 14 (1.9%) | 1 (0.5%) | 3 (2.0%) | 0 | 2 (13.3%) |
| Other (%) | 11 (1.5%) | 3 (1.5%) | 1 (0.7%) | 0 | - |
| Respondent's Relationship to Child (%) |  |  |  |  |  |
| Child: School Attendance Problems | 100% | 100% | 0% | n/a | 18.2% |
| Child: Neurodivergence | 666 (92.1%) | 168 (83.6%) | 33 (22.2%) | 22 (88.0%) | 4 (36.4%) |
| Child: Autism | 598 (83.4%) | 133 (66.2%) | 25 (16.8%) | 13 (52.0%) | 2 (10.6%) |
| Child: Sensory Processing Difficulties | 406 (56.9%) | 87 (43.3%) | 10 (6.7%) | 13 (52.0%) | 1 (5.3%) |
| Child: ADHD | 395 (55.3%) | 87 (43.3%) | 13 (8.7%) | 12 (48.0%) | 2 (10.6%) |
| Child: Mean number of ND (StDev) | 3.7 (2.5) | 3 (2.5) | 0.72 (1.7) | 2.5 (2.0) | 1 (0) |
| Parent/Professional: Neurodivergence: |  |  |  |  |  |
| Yes | 150 (21.3%) | 47 (24.6%) | 10 (6.7%) | 5 (21.7%) | 1 (7.14%) |
| Maybe | 338 (48.0%) | 72 (37.7%) | 23 (15.4%) | 10 (43.5%) | 1 (7.14%) |
| No | 216 (30.7%) | 72 (37.7%) | 116 (77.9%) | 8 (34.8%) | 13 (92.86%) |

#### Professionals

Professionals due to attend a conference on school anxiety in the North of England were invited to participate. 19 professionals participated (mean completion rate = 82.84%, with 78.95% completing 100%). Their roles included teachers, teaching assistants, higher level teaching assistants, SENCOs, headteachers, deputy headteachers, school nurses, attendance inclusion officers, welfare managers, child psychologists/psychotherapists, and educational psychologists. Most participants were also mothers, and four were parents of neurodivergent CYP (Table 1). One participant identified as neurodivergent. Participants had a range of experience working with CYP accessing education in different educational provisions and/or without educational provision for a variety of reasons (Table S1) and ranged in confidence when supporting CYP with school attendance problems and/or autistic CYP (Table S2).

### 2.2. Research Ethics and Language

This study was approved by the Faculty of Medical Sciences Research Ethics Committee, part of Newcastle University’s Research Ethics Committee. Where possible, we use identity-first language (e.g., autistic CYP) (23). We defined neurodivergence (ND) for parents as “a term for when someone’s brain processes, learns and behaves differently from what is considered ‘typical’. Autism is an example of a neurodivergence.” We use the term ‘neurotypical’ (NT) to refer to CYP whose parent identified them as not being neurodivergent. When designing and conducting this study, we used the term ’school-refusal’ to refer to CYP unable to attend school due to the emotional distress experienced at school. During the data analysis, it became evident that this was not an appropriate terminology, at which point we coined the term School Distress to describe this experience more appropriately and to ensure the CYP’s experience is front and centre stage of discussion (5).

### 2.3. Design

The study employed a case-control, concurrent embedded mixed-methods design, within which qualitative data was collected to supplement quantitative data. This was chosen due to the exploratory nature of this study, and because the limited literature base prevented us from providing fully comprehensive lists of response options to some questions. To collect qualitative data, free text boxes were presented within some questions for parents to provide additional comments.

### 2.4. Materials

Whilst the full questionnaire used here is described elsewhere (5), questions relating to the parent/professional lived experience of School Distress are described here. These are considered under five related categories: (1) the direct impact of supporting a child or young person experiencing School Distress on the parent/professional themselves, (2) the interactions that parents and professionals have with others, including the professionals/services surrounding the child and family, and wider family, friends, and acquaintances, (3) actions taken against parents to enforce school attendance and the impact of these actions, (4) perspectives with respect to underlying drivers of school attendance problems, and (5) the key sources of support available to parents of CYP experiencing School Distress.

#### 2.4.1. Direct Impact of Supporting a Child with School Distress

##### 2.4.1.1. Impact on Mental Health

###### Mood and anxiety levels

All participants were asked to quantify their typical daily mood (0=very negative to 10=very positive) and the level of daily anxiety they currently experience (0=none to 10=high). Parents of CYP currently experiencing School Distress were also asked to rate their typical daily mood and anxiety prior to the onset of their child’s school attendance problems. Parents who rated their children’s school attendance problems as historical (Past SD) were asked to quantify their typical daily mood and anxiety both before and during their child’s School Distress, as well as currently.

###### Discrete Emotions Questionnaire

To comprehensively describe the emotional lived experience of supporting a CYP with School Distress, all participants were also asked to complete the Discrete Emotions Questionnaire (24). This self-report scale consists of 32 items, aiming to measure eight distinct state emotions, with 4 emotions per emotion state (see Figure 3A). Individuals respond along a 7-point Likert scale (1=not at all, 7=an extreme amount), stating the extent to which they experience each of the 32 items. Total scores are then calculated by summing participant’s responses to each subscale. This scale has excellent internal consistency (α>0.82 for each subscale).

School Distress parents responded based on their emotions when their child was experiencing School Distress. Parents of Lifelong EHE CYP and participants in the professional group responded with respect to a period of time of at least a few months over the last year (excluding Covid-19 lockdowns), and parents of children who do not experience School Distress (No SD) were asked to think about an equivalent period of time over the last year where their child was attending school.

###### New Mental Health Condition

Finally, participants who had parented a child with School Distress (Current or Past) were also asked whether they had developed a new mental health condition since the onset of their child’s school attendance difficulties.

##### 2.4.1.2. Wider Impact of School Attendance Problems

Using a Likert scale, parents in the two School Distress groups (Current and Past SD) rated the impact that supporting a CYP with School Distress has on their own physical health, relationships, career, and financial situation, as well as the impact on their other children, wider family, and family friends (0=no negative impact, 5=some negative impact, 10=considerable negative impact). Professionals who reported having direct experience working with a child or children with school attendance difficulties also completed this section – with the question being whether the experience of working with children with school attendance difficulties, and the associated events, had negatively impacted their own physical health, their relationships, their career, their financial situation, their other children, their wider family, and their family friends.

All participants were provided the opportunity to discuss ’Other’ impacts.

##### 2.4.1.3. School Distress as a Threatening Life Event

To understand how the experience of parenting a child with School Distress compares to other stressful or threatening life events, we utilised the List of Threatening Life Experiences (LTE) (25). This is a list of twelve life event categories with considerable long-term contextual threat, including ’serious illness or injury to self’, ’death of a first-degree relative, including spouse or child’, and ’major financial crisis’ (see Supplementary Notes 2). The LTE has high test-retest reliability, good agreement with informant information, and both high specificity and sensitivity (26). The LTE has also been found to have good validity and stability over time (27).

Here, we adapted the LTE to include the original 12 threatening life events, plus 6 additional life events. Five of the additional life events were taken from Burghal et al.’s Appendix B, which presents a list of 15 prescribed life events considered to have mild or no long-term threat (i.e. ’had a baby’, ’a minor injury or illness to self’, ’started a different type of job’, ’had moderate financial difficulties’, and ’moved house within own town/city’) (25), whilst the sixth was ’child school-refusing’.

All groups were asked to select what they considered to be the top 10 most stressful life events from this list of 18 categories. Participants were then asked to sort their selected 10 life events in order, starting with that which they considered to be the most threatening and ending with that which they considered to be the least threatening. Participants were reminded that they did not need to have experienced all events personally to rank them.

For scoring and analysis purposes, the most threatening life event selected by each participant was given a score of ‘10’, with the second most threatening scored ‘9’. This proceeded until the item ranked lowest within the selected 10 life events was scored ‘1’, after which all life events not selected within the top 10 were scored as ‘0’. These scores were used to compute an average score for each of the 18 items for each of the five participant groups respectively.

#### 2.4.2 Interactions with Individuals Surrounding Child and Family

##### 2.4.2.1 Tone of Communication used by Professionals

Parents were presented with a list of 27 adjectives (’Adversarial’, ’Aggressive’, ’Calm’, ’Caring’, ’Compassionate’, ’Conspiratorial’, ’Critical’, ’Disrespectful’, ’Dismissive’, ’Friendly’, ’Guarded’, ’Helpful’, ’Hostile’, ’Hurtful’, ’Informed’, ’Intimidating’, ’Kind’, ’Optimistic’, ’Respectful’, ’Unclear’, ’Understanding’, ’Uninformed’, ’Unsupportive’, ’Secretive’, ’Supportive’, ’Sympathetic’, ’Threatening’), plus an ’Other’ option (including a free-text box for participants to enter the appropriate adjective). All parents were asked to select which adjectives they felt appropriately described the tone of communication used by professionals when communicating with them. Professionals in this context was defined for participants as being “anyone who is working in a professional (e.g., paid) capacity with your child (e.g., health care professionals, children’s social services, local authority EHE staff…etc.)”.

##### 2.4.2.2 Not Feeling Believed

Parents were asked whether they have ever felt like they have not been believed when they have raised concerns about their child’s difficulties [response options: ’No’, ’Yes, by school staff’, ’Yes, by health care professionals’, and ’Yes, by others (please specify)’]. Professionals were asked whether they have ever felt like they have not been believed when they have tried to raise concerns about a child’s difficulties [response options: ’No’, ’Yes, by the CYP’s parents’, Yes, by school staff’, ’Yes, by health care professionals’, and ’Yes, by others (please specify)’].

##### 2.4.2.3 Experience of Professional Gaslighting

All parents were asked if they have ever experienced professional gaslighting (defined as an interaction “where a professional makes you question your own reality”). Three response options were provided: ’No, never’, ’Yes, occasionally’, and ’Yes, frequently’. Professionals were not asked this question.

##### 2.4.2.4 Feeling Threatened or Vulnerable

All parents were asked whether, as a parent/carer, they have ever felt threatened or vulnerable as a result of an interaction with a member of school staff (response options: ’No’, ’Unsure’, and ’Yes, definitely’). As with above, the professional group was not asked this question.

#### 2.4.3 Action Taken Against Parents to Enforce Attendance

Parents in the Current SD and Past SD groups were asked whether they have ever had any action taken against them to enforce their child’s school attendance [response options: ’No’, ’Yes, a fine (sometimes known as a ‘penalty notice’)’, ’Yes, a Parenting Order’, ’Yes, an Education Supervision Order’, ’Yes, a School Attendance Order’, ’Yes, I was prosecuted and given a Community Order’, ’Yes, I was prosecuted and given a jail sentence’, ’Yes, a Fabricated or induced illness (FII) accusation’, ’Yes, Child Protection Procedures’, and ’Other (please provide details below)’]. Parents were asked to select all options that applied to their situation.

Professionals were also asked whether they (or a colleague working with them on a case) had ever taken any of the above actions against a parent(s) to enforce school attendance. They were also asked whether they had ever punished or rewarded a CYP personally because of their school attendance record.

#### 2.4.4 Causal Factions

Following a comprehensive, collaborative review of the literature, encompassing a multitude of factors which have been suggested to underpin school attendance problems, we compiled a list of 98 potentially causal factors of School Distress. This included an ‘other’ item with a free text box. For clarity of presentation here, these 98 items have been classified into 12 broad categories i.e., ‘Mental Health’ (containing 7 items), ‘Physical Health’ (3 items), ‘Worries/Negative Emotions’ (11 items), ‘School-Related Factors’ (22 items), ‘Academic Factors’ (3 items), ‘Disability-Related Factors’ (15 items), ‘Peer Relations’ (9 items), ‘Pupil Behaviour’ (3 items), ‘Teacher-Related Factors’ (6 items), ‘Reward/Punishment by School Staff’ (4 items), ‘Parent/Family-Related Factors’ (9 items), and ‘Other’ (6 items). For complete list, see Supplementary Notes 4/5.

School Distress parents were asked to identify the reasons underlying their child’s difficulties attending school from this list. Control parents and professionals were asked to select factors which they believed may be the reasons that children experience school attendance difficulties. Once participants had identified what they believe to be causal factors, they were then asked to identify the most important factor(s). Participants were instructed to limit this selection to a maximum of 3 factors.

The rationale for including responses to this question within this paper, is that it permitted a statistical exploration of differences in opinion with respect to the most important drivers of School Distress from a parental lived-experience perspective relative to the professional perspective. Hence, the data from control parents is not included here.

#### 2.4.5 Sources of Support for Parents and Professionals

Finally, using a free text box, all parents were asked: “As a parent, what has been your most important source(s) of support?”, whereas professionals were asked “As a professional working with CYP with school attendance difficulties, what has been your most important source(s) of support?”. Professionals were also asked “In your experience, how do you believe professional support could and/or should be improved, and how would this have benefitted you personally, and the CYP you were supporting?”. Professionals were also asked whether they would like a) more support and b) more training ‘When supporting CYP with School Attendance Problems’ and ‘When supporting Autistic CYP’.

### 2.5. Procedure

Data was collected using Qualtrics. The parent survey link was shared widely on social media, and the additional control participants recruited via prolific.org were directed to the Qualtrics link. Participants in the professional group were attendees at a conference on school anxiety and were invited to participate prior to attending the conference. Invitations were sent via email by the conference organisers. After reading the information sheet, participants provided written consent before commencing. Participants were informed that they could skip any questions and stop/start at any time. Qualtrics’ display-logic function ensured respondents were only asked questions which were relevant to them. Upon completion, participants were presented with a debrief form, including a comprehensive list of support services. The parent study ran for 14 days (22/02/2022–08/03/2022) and the professional study was conducted in January 2023.

### 2.6. Data Analysis

Quantitative data analyses were run using IBM SPSS Statistics V26. Descriptive statistics were calculated to summarise participants’ responses to each question. Further statistical analyses were then conducted to examine relationships between variables. Before performing statistical analyses, Normality was assessed by plotting results in histograms and conducting Shapiro-Wilk and Kolmogorov-Smirnov tests. When results were not normally distributed, non-parametric methods were used (e.g., Kruskal-Wallis tests with Mann-Whitney U post-hoc analyses examined differences in anxiety and mood scores between School Distress groups). Chi-squared tests were used to determine whether there was a statistically significant difference between the expected frequencies and the observed frequencies in one or more categories. A significance level of α=0.05 was adopted for all analyses, except during post-hoc tests where Bonferroni adjustment was applied.

Qualitative analysis was used to analyse additional comments provided by parents in response to some survey questions. In the interest of space, a thematic analysis of just one question is reported here (i.e., free text comments in response to the question “Have you ever had any action taken against you to enforce school attendance?”). This question was chosen as, in this instance, the ’Other’ option was the second most endorsed option (after ’No’), with 106 parents providing a free text comment. The volume of responses indicated value in formally analysing these free-text comments.

Qualitative data analysis followed the six phases of thematic analysis recommended by Braun and Clarke (28), aiming to identify key themes within the data to help answer our research question. During analysis, an inductive approach was taken, such that codes and themes were developed from the content of the dataset itself, rather than any prior theoretical commitments. Given the current lack of in-depth research into the experience of School Distress, this enabled us to identify new, valuable information. An essentialist/realist position was taken, assuming a unidirectional relationship between the participants’ experiences and their language used.

## 3. Results

### 3.1. Direct Impact of Supporting a Child with School Distress

#### 3.1.1. Impact on Mental Health

##### Mood and anxiety levels

Current mood differed significantly between groups (Kruskal-Wallis test: *p*<.001), with post-hoc Mann-Whitney U tests indicating that mood levels were significantly lower in parents in the Current SD group compared to all of the other four groups (i.e., the Past SD group, No SD group, Lifelong EHE group and Professional group). Current anxiety levels also differed between groups (*p*<0.001), with parents of children currently experiencing School Distress reporting significantly greater levels of daily anxiety than parents in the other three parent groups (i.e., the Past SD, No SD, and Lifelong EHE groups) and the Professional group (see Figure 1 for full details).

**Figure 1:**
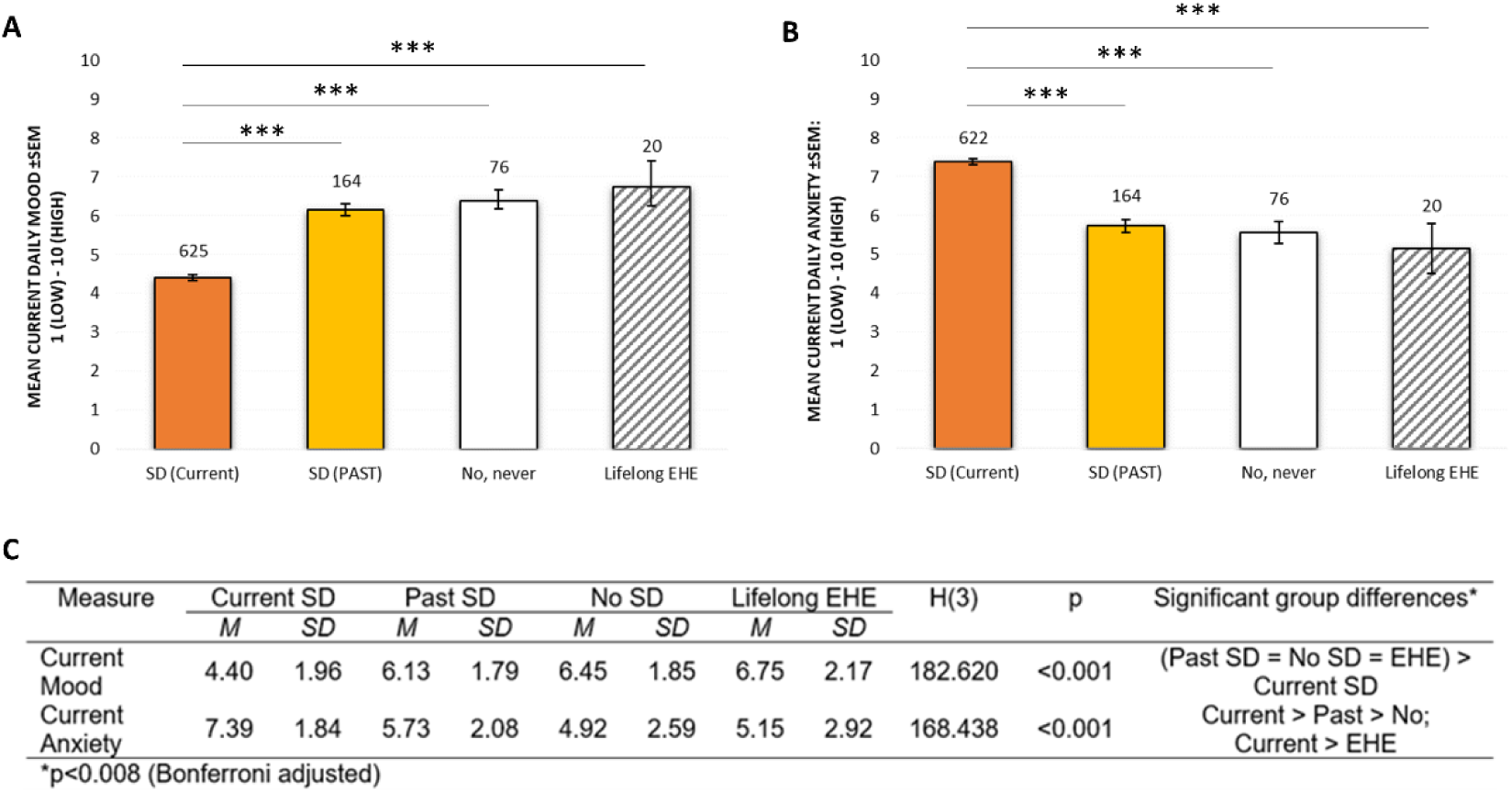
**Panel A.** Mean Mood and Anxiety Levels currently experienced by respondents in each of the five groups. Error bars represent +/- 1 SEM. **Panel B.** Details the results of the between-group Kruskal-Wallis analyses and subsequent post-hoc Mann-Whitney U tests.

In addition to providing estimates of current mood and anxiety, parents in the Current SD group also provided retrospective estimates of their mood and daily anxiety levels before their child’s School Distress began. Parents reported significantly higher mood and significantly lower daily anxiety before their child’s School Distress commenced (see Figure 2A).

**Figure 2:**
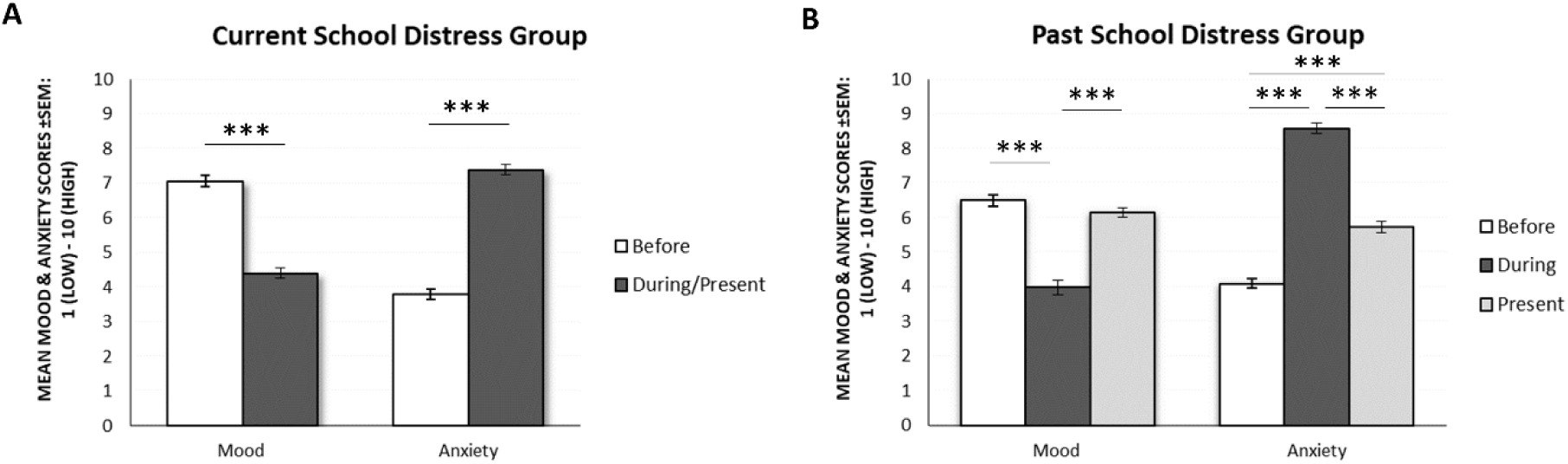
**Panel A - Current School Distress Group**: Mean mood and anxiety levels of parents in the Current School Distress before and during their child’s School Distress. **Panel B - Past School Distress Group**: Mean mood and anxiety levels of parents in the Past School Distress before, during and after their child’s School Distress. Error bars represent +/- 1 SEM, *** *p* < .001.

**Figure 3:**
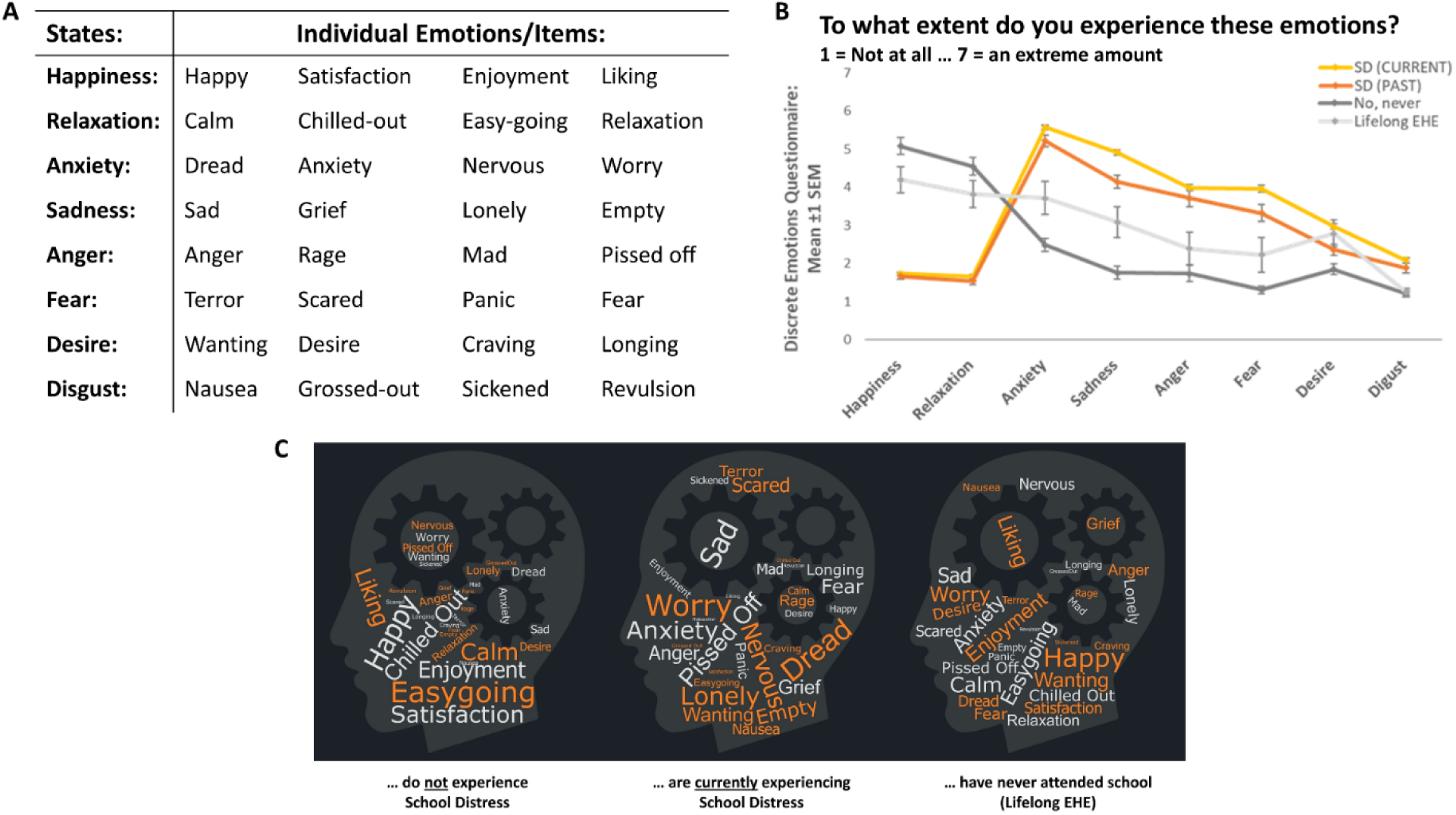
**Panel A.** The 8 emotion states measured by the DEQ (left-hand column). Each emotion state is a composite score of four related emotions. These individual, related emotions are listed after each emotion state. **Panel B.** The mean ratings provided by each parent group for each of the eight emotion states. Error bars represent +/- 1 SEM. **Panel** C. Word Clouds illustrate the parent ratings of each of the 32 individual emotions assessed in the DEQ. The size of the word in the clouds represents how strongly each emotion was experienced by parents within the No SD control Group, the Current SD group, and the Lifelong EHE parent group. The bigger the word, the more strongly it was endorsed by parents within each group. Word clouds were generated using https://www.wordclouds.com/

Similarly, parents whose child’s School Distress was historical (i.e., parents in the Past SD group) also retrospectively estimated their mood and daily anxiety levels before their child’s School Distress began, in addition to providing retrospective estimates of their mood and daily anxiety levels during their child’s School Distress. Importantly, the change in parental mood and anxiety pre-School Distress relative to during-School Distress in the Past SD group mirrored that reported in the Current SD group (see Figure 2B). Reassuringly, the Past SD group also showed a relative recovery of their mood and daily anxiety levels once their children’s School Distress resolved (often via parents removing their children from a school setting and educating them at home themselves), although parental anxiety levels post-School Distress did remain significantly higher than pre-School Distress (Figure 2B).

##### Discrete Emotions Questionnaire (DEQ)

A score for each of the eight distinct emotion states (i.e., anger, disgust, fear, anxiety, sadness, happiness, relaxation, and desire) was computed from the 32 individual emotions sampled in the DEQ. Significant between-group differences were evident in each (see Figure 3B). Mann-Whitney U post-hoc tests revealed that during their child’s School Distress, parents in the Current and Past SD groups experienced significantly lower levels of positive emotion states (relaxation and happiness) and significantly higher levels of all negative emotion states (anger, anxiety, sadness, disgust, and fear) relative to the parents in the No SD and Lifelong EHE groups, and to the professionals (see Table 2).

**Table 2:**
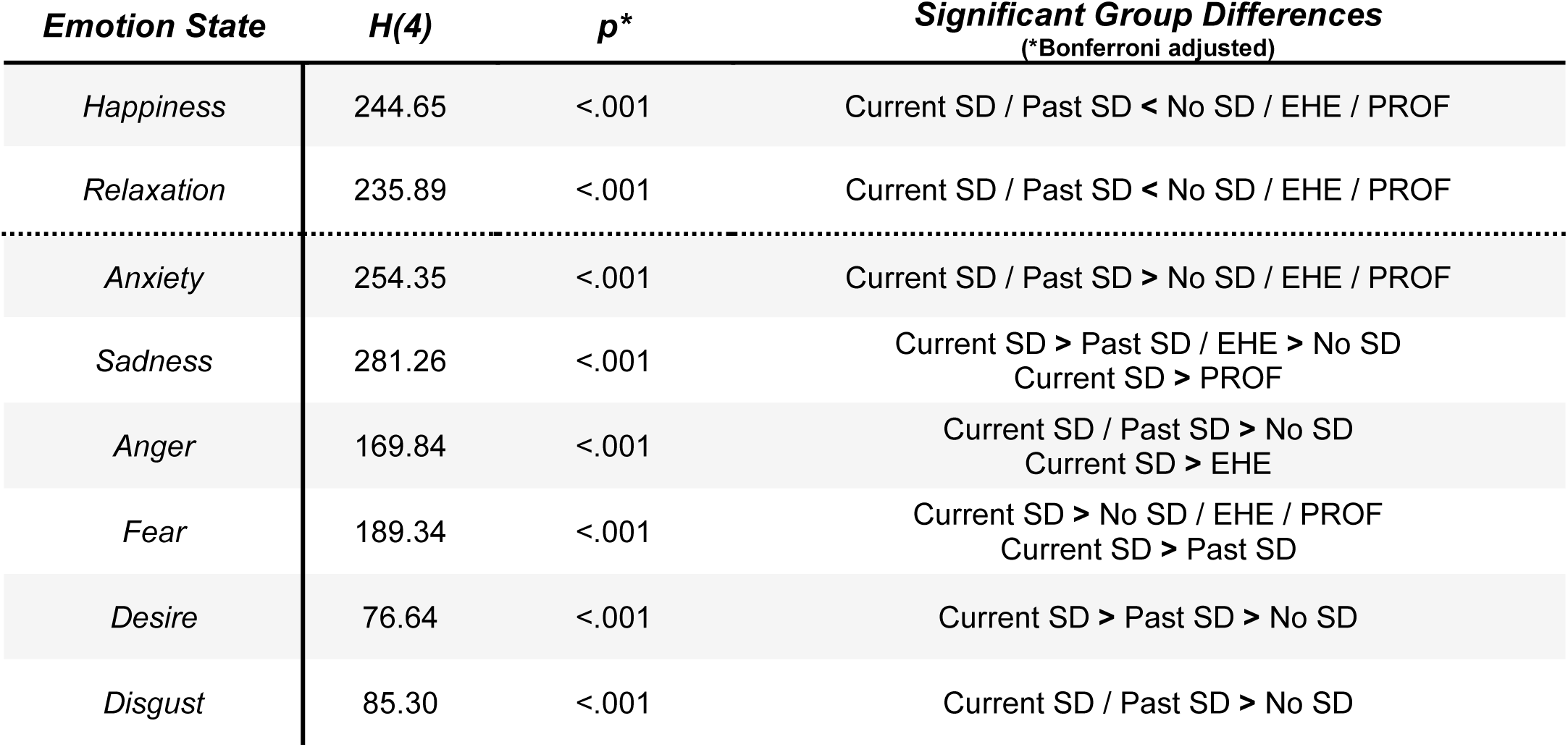
Significant Group Differences in DEQ Scores – Kruskal Wallis H test with Mann-Whitney U post-hoc tests (Bonferroni adjusted). SD – School Distress, EHE – Lifelong Electively Home-Educated, PROF = Professionals.

| *Emotion State* | *H(4)* | *p\** | *Significant Group Differences*<br>(*Bonferroni adjusted) |
| --- | --- | --- | --- |
| *Happiness* | 244.65 | <.001 | Current SD / Past SD < No SD / EHE / PROF |
| *Relaxation* | 235.89 | <.001 | Current SD / Past SD < No SD / EHE / PROF |
| *Anxiety* | 254.35 | <.001 | Current SD / Past SD > No SD / EHE / PROF |
| *Sadness* | 281.26 | <.001 | Current SD > Past SD / EHE > No SD<br>Current SD > PROF |
| *Anger* | 169.84 | <.001 | Current SD / Past SD > No SD<br>Current SD > EHE |
| *Fear* | 189.34 | <.001 | Current SD > No SD / EHE / PROF<br>Current SD > Past SD |
| *Desire* | 76.64 | <.001 | Current SD > Past SD > No SD |
| *Disgust* | 85.30 | <.001 | Current SD / Past SD > No SD |

Parental responses to each of the 32 individual emotions in the DEQ are represented at the group level qualitatively in Figure 3C. The larger the word in these word clouds, the more strongly this emotion was endorsed by the parents in the respective groups (i.e., No SD group, Current SD group, Lifelong EHE group).

##### Development of new mental health condition

51.7% of parents in the Current SD group, and 42% of parents in the Past SD group, reported having developed a new mental health condition (diagnosed or suspected) since their child’s School Distress began.

#### 3.1.2. Wider Impact

One-sample t-tests (where no impact = 0) found that School Distress had a significant, negative impact on every aspect of the parents’ lives measured - on parental physical health, their careers, their financial situation, their other children, their wider family unit and family friends, and on their relationships with their partners (see Figure 4 and Supplementary Note 3). Both the Current and Past SD groups reported the most negative impact as being on their own careers, followed closely by their other children, their financial situation, and their relationship with their partner. When parents reported ’Other’ negative impacts, free text comments indicated that this most frequently referred to the deleterious impact on their own mental health (see Discussion for further descriptions). No significant negative impact on professionals’ lives was found (all *p*’s > .05) (see Figure 4).

**Figure 4:**
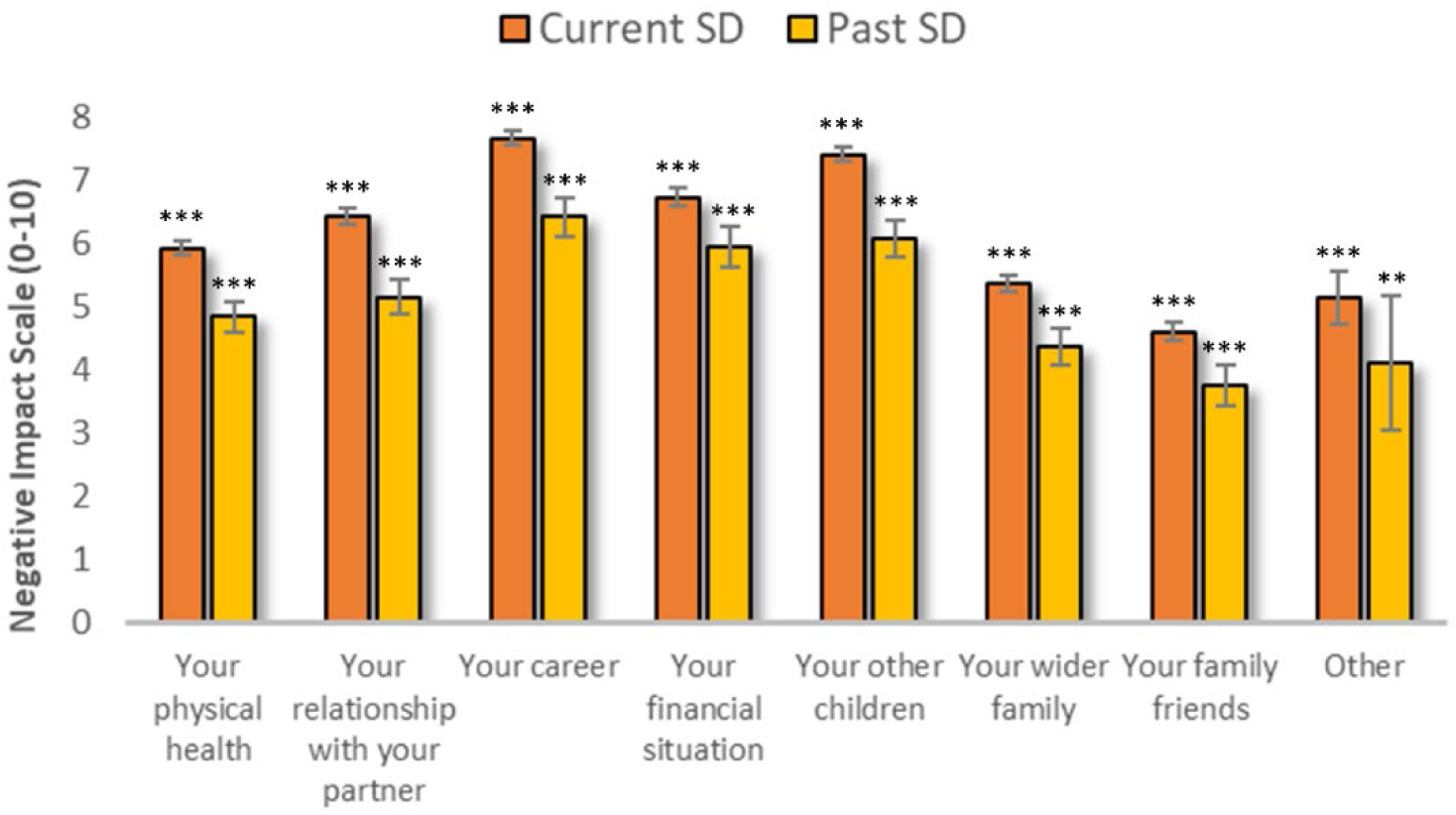
The Mean Extent to Which School Distress Has a Wider Negative Impact on the Respondent’s Life. Error bars represent +/- 1 SE. Note: Responses were rated on a scale of 0-10, where 0 indicates no negative effect and 10 indicates a very large negative effect. One-sample t-tests revealed that the mean scores for each variable were significantly greater than 0: i.e., there was a significant negative impact of School Distress on parents’ (1) physical health [Current SD: t(526) = 49.15, *p* < .001; Past SD: t(127) = 19.22, *p* < .001], (2) their relationships with their partners [Current SD: t(460) = 49.01, *p* < .001; Past SD: t(119) = 18.28, *p* < .001], (3) their careers [Current SD: t(509) = 67.94, *p* < .001; Past SD: t(121) = 21.67, *p* < .001], (4) their financial situation [Current SD: t(476) = 48.58, *p* < .001; Past SD: t(121) = 18.43, *p* < .001], (5) their other children [Current SD: t(466) = 66.66, *p* < .001; Past SD: t(102) = 20.55, *p* < .001], (6) their wider family unit [Current SD: t(476) = 42.34, *p* < .001; Past SD: t(114) = 14.66, *p* < .001], (7) their family friends [Current SD: t(413) = 29.87, *p* < .001; Past SD: t(97) = 11.66, *p* < .001], and on (8) ’other’ [Current SD: t(97) = 12.26, *p* < .001; Past SD: t(18) = 3.85, *p* < .01]. Error bars represent +/- 1 SEM. * *p* < .05, ** p < .01, *** *p* < .001.

#### 3.1.3. School “Refusal” as a Threatening Life Event

Parents in the No SD (control) group perceived the experience of a ‘Child School Refusing’ as the 10th most threatening life event, relative to the 9 other events that they selected to be most threatening (see Table 3). Parents whose children have never attended school (Lifelong EHE) also placed the experience of having a ‘Child School Refusing’ low with respect to other threatening life events, with it not appearing at all within their selection of the top 10 most threatening life events. Instead, this experience fell in joint 12th place with the experience of ‘having moderate financial difficulties’.

**Table 3:** Most to Least Threatening Life Events, as by participants in each of the five groups. Events are ordered based upon the mean position they were placed by participants. Note. Events separated by ; were chosen as equally threatening.

| Most to Least Threatening | Current SD | Past SD | No SD | Lifelong EHE | Professionals |
| --- | --- | --- | --- | --- | --- |
| 1st | Death of a 1st degree relative including child or spouse | Death of a 1st degree relative including child or spouse | Death of a 1st degree relative including child or spouse | Death of a 1st degree relative including child or spouse | Death of a 1st degree relative including child or spouse |
| 2nd | **Child "School Refusing"** | Serious Illness or Injury to Self; Death of close family friend or 2nd degree relative | Serious Illness or Injury to Close Relative | Serious Illness or Injury to Self | Serious Illness or Injury to Self |
| 3rd | Serious Illness or Injury to Self | Major financial crisis | Serious Illness or Injury to Self | Serious Illness or Injury to Close Relative | Death of close family friend or 2nd degree relative |
| 4th | Serious Illness or Injury to Close Relative | **Child "School Refusing"** | Death of close family friend or 2nd degree relative | Major financial crisis | **Child "School-Refusing"** |
| 5th | Death of close family friend or 2nd degree relative | Serious Illness or Injury to Close Relative | Major financial crisis | Death of close family friend or 2nd degree relative | Separation due to marital difficulties |
| 6th | Major financial crisis | Separation due to marital difficulties | Separation due to marital difficulties | Separation due to marital difficulties | Major financial crisis |
| 7th | Separation due to marital difficulties | Problems with police/court appearance | Sacked from job | Serious problem with close friend, neighbour or relative; Sacked from job | Serious Illness or Injury to Close Relative |
| 8th | Problems with police/court appearance | Sacked from job | Problems with police/court appearance | Had a baby | Problems with police/court appearance |
| 9th | Sacked from job | Had a baby | **Child "School-Refusing"** | Problems with police/court appearance | Sacked from job |
| 10th | Serious problem with<br>close friend,<br>neighbour or relative | Serious problem with<br>close friend, neighbour<br>or relative;<br>Unemployed seeking<br>work for more than 1<br>month | Unemployed seeking<br>work for more than 1<br>month | Unemployed seeking<br>work for more than 1<br>month | Had a baby |

In contrast, parents of CYP currently experiencing School Distress collectively rated a ‘Child School Refusing’ as the 2nd most threatening life event, only superseded by the ’Death of a 1st degree relative including child or spouse’. Parents with historical School Distress experience (Past SD) collectively rated this experience as the 5th most threatening life event category, superseded by ‘Death of a 1st degree relative including child or spouse’, ‘Serious Illness or Injury to Self’, ‘Death of close family friend or 2nd degree relative’ and ‘Major financial crisis’.

The Professional group rated the experience of a ‘Child School Refusing’ as the 4^th^ most threatening life event, superseded by ‘Death of a 1st degree relative including child or spouse’, ‘Serious Illness or Injury to Self’, and ‘Death of close family friend or 2nd degree relative’. Thus, qualitatively, this group showed an understanding more in keeping with parents with direct experience of school attendance problems than parents without.

A Kruskal-Wallis test was performed on the scores of the five groups (see Figure 5). With respect to scores for a child “school refusing”, the differences between the rank were significant, H (4, n=841) = 170.57, *p*<.001. Post-hoc comparisons conducted using Mann-Whitney U Tests (with a Bonferroni adjusted alpha level) revealed that parents in both the Current and Past SD groups rated the experience of a child “school refusing” significantly more threatening than control parents (i.e., the No SD and Lifelong EHE groups; *p*’s < .001, adjusted sig.). Similarly, the professional group rated the experience of a child “school refusing” as significantly more threatening than control parents (i.e., the No SD and Lifelong EHE groups; *p*’s < .05, adjusted sig.); with no significant differences between SD parent groups and the professional group (*p*’s = 1.0, adjusted sig.). Finally, parents in the Current SD group rated the experience of a child “school refusing” as significantly more threatening than the Past SD group (*p*<.05, adjusted sig.).

**Figure 5:**
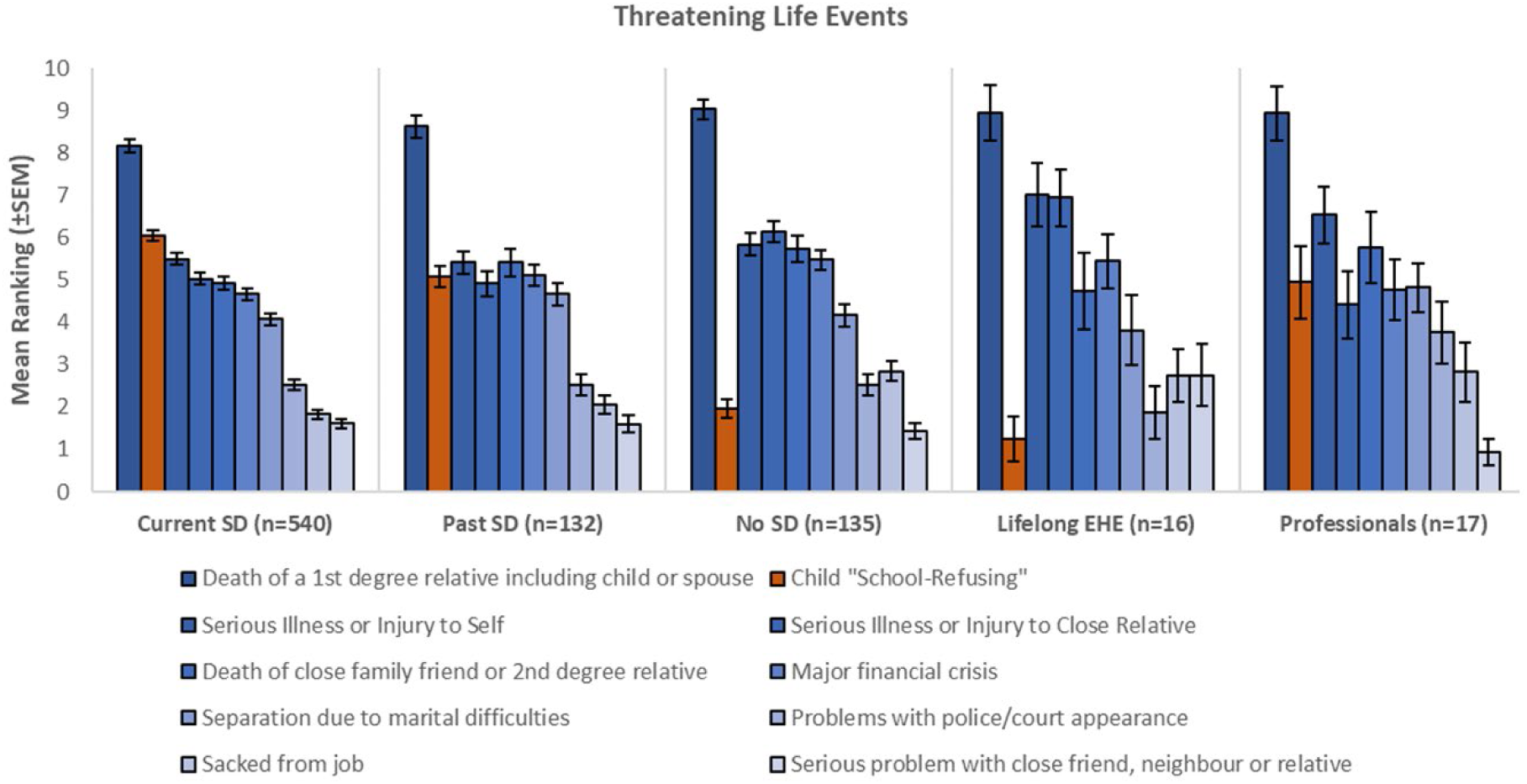
Threatening Life Events. Current SD grouping: Mean rankings for the top 10 most threatening life events selected by the parents in the Current SD group. The mean rankings for these 10 life events provided by participants in the other 4 groups follow to the right, with the mean rankings for the experience of having a child ‘school refuse’ represented in brown. Error bars represent +/- 1 SEM.

### 3.2. Interactions with Individuals Surrounding Child and Family

#### 3.2.1. Tone of Communication

When asked to identify adjectives that best describe the tone of communication used by professionals when communicating with them, parents of children with experience of School Distress most frequently selected negative adjectives, such as ‘dismissive’, ‘critical’, ‘unsupportive’, and ’uninformed’. Some positive adjectives were selected, but less frequently than the above negative emotions (see Table 4). This was consistent in both the Current SD and Past SD groups, and contrasted with the parents of children who have never experienced School Distress (i.e., the No SD group), who most frequently selected positive adjectives such as ‘friendly’, ‘calm’, ’caring’ and ’helpful’.

**Table 4:** Most frequently selective adjectives by parents to describe the tone of communication used by professionals when communicating with them. Note: for parents in both the Current SD and Past SD groups, this referred to communications specifically during their child’s School Distress.

| Position (Most to Least Commonly Used) | Current SD | Past SD | No SD | Lifelong EHE |
| --- | --- | --- | --- | --- |
| 1st | Dismissive | Dismissive | Friendly | Caring/<br>Friendly |
| 2nd | Critical/<br>Unsupportive | Unsupportive | Calm | Helpful |
| 3rd | Uninformed | Uninformed | Helpful | Calm |
| 4th | Calm | Critical/Caring | Respectful | Compassionate<br>/ Respectful |
| 5th | Caring | Compassionate/<br>Friendly | Caring | Kind /<br>Uninformed |

Parents were also able to add additional adjectives into a free text box. Parents with experience of School Distress included adjectives such as ‘arrogant’, ‘apologetic’, ‘cold’, ‘condescending’, ‘confusing’, ‘deceitful/lying’, ‘derogatory’, ‘disbelieving’, ‘disempowering’, ‘duplicitous’, ‘friendly’, ‘gaslit’, ‘helpful’, ‘ignorant’, ‘inconsistent’, ‘irritated’, ‘negative’, ‘nice’, ‘respectful’, ‘underhand’, ‘unprofessional’, ‘unsure’, ‘pacify’, ‘patronising’, ‘sarcasm’, ‘scary’, ‘sceptical’, ‘trivialising’, ‘voiceless’, and ‘wrong’. Of the additional adjectives, ‘patronising’, ‘deceitful/lying’, ‘condescending’ and ‘sarcasm’ were reported most frequently.

#### 3.2.2. Not Feeling Believed

##### Not feeling believed by school staff/parents

When directly asked if they had ever felt like they have not been believed when they have raised concerns about their child’s difficulties, 78.7% of parents in the Current SD group and 75.7% of parents in the Past SD group reported that they have felt this way, compared to just 17.8% of parents in the No SD group (see Figure 6A). A chi-square test was performed to examine the relationship between feeling believed by school staff and being a parent with or without experience of School Distress (Current SD/Past SD versus No SD). School Distress parents were significantly more likely to report not being believed by school staff than control (No SD) parents (X^2^ (1, N = 658) = 160.34, *p* < .001). In addition, 28.6% of participants in the Professional group also reported not feeling believed by school staff when raising concerns about a child’s difficulties. This percentage included a SENCO, a senior psychological wellbeing practitioner, a pastoral manager, and an attendance inclusion officer. However, only 10.5% of participants in the Professional group reported not being believed by the CYP’s parents when raising concerns with them about a child’s difficulties.

**Figure 6:**
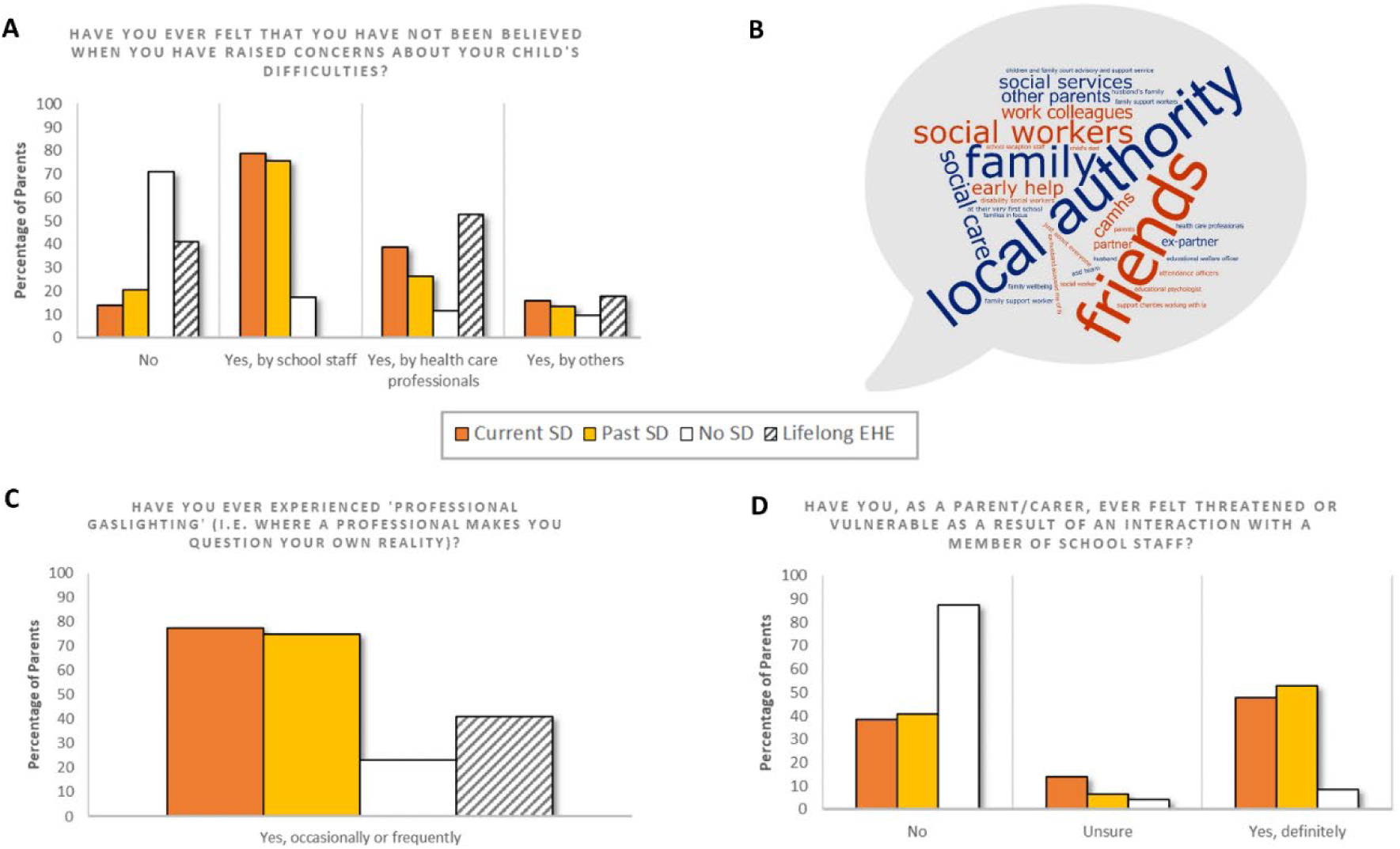
**Panel A**. Percentage of participants who have ever felt as if they have not been believed by School Staff, Health Care Professionals, or ’Others’, when they have raised concerns about their/a child’s difficulties. **Panel B.** A word cloud of the free text responses provided by parents to the question of who else (i.e. the ’Others’ in Panel A) did not believe them when they raised concerns about their child’s difficulties. The bigger the word, the more frequently this option was disclosed by parents. **Panel C.** Percentage of parents who reported ’never’, ’occasionally’, and ’frequently’ experiencing professional gaslighting i.e., where a professional made them question their own reality. **Panel D.** Percentage of parents who have ever felt threatened or vulnerable because of an interaction with a member of school staff.

##### Not feeling believed by health care professionals

38.7% of parents in the Current SD group and 26.2% of parents in the Past SD group reported having felt disbelieved by health care professionals when they have tried to raise concerns about their child’s difficulties, relative to only 10.2% of parents in the No SD group and 5.3% of professionals. As with school staff, parents with experience of School Distress were significantly more likely to report not being believed by health care professionals when raising concerns about their child’s difficulties than those with no experience of School Distress [X^2^ (1, N = 658) = 30.58 *p* < .001].

Notably, 52.9% of parents in the Lifelong EHE group reported that they have felt disbelieved by health care professionals previously. This was significantly higher than in the No SD control group [X^2^ (1, N = 135) = 20.69, *p* < .001] and Past SD group [X^2^ (1, N = 120) = 4.64, *p* < .05], but did not differ significantly relative to the Current SD group (X^2^ (1, N = 454) = 1.39, *p* = .237).

##### Not feeling believed by others

In addition, many parents indicated that they felt that they have not been believed about their child’s difficulties ’by others’, with the majority of these parents falling into the School Distress (Current and Past) or Lifelong EHE groups. A total of 84 parents provided free text comments with respect to who they were referring to. Responses are represented in Figure 6B. Children’s Social Services (x18), family (x14), friends (x11) and Local Authorities (x10) were the most frequently mentioned by parents (although different descriptors were used for Children’s Social Services: 5 x Social Workers, 4 x Social Care, 3 x Social Services, 3 x Early Help, 1 x Disability Social Worker, 2 x Family Support Worker). In addition, partners, ex-partners, and ex-husbands were mentioned by several parents, as well as other parents, work colleagues, and the parent’s own parents. More specialist mental health services, such as CAMHS, educational psychology, and an ASD-team were also mentioned.

#### 3.2.3. Experience of Professional Gaslighting

77.6% of parents of children currently experiencing School Distress, and 69.9% of parents of children who have experienced School Distress in the past, reported either occasionally or frequently experiencing professional gaslighting (see Figure 6C), which is where individuals are manipulated “into doubting his or her perceptions, experiences, or understanding of events” (American Psychological Association, n.d.). A Kruskal-Wallis H test with Mann-Whitney U post-hoc analyses revealed that significantly more parents in the Current and Past SD groups reported either occasionally or frequently experiencing gaslighting compared to parents in the No SD group ([Current SD/Past SD] > No SD; Current SD > [Lifelong EHE]).

#### 3.2.4. Feeling Threatened or Vulnerable

Furthermore, 47.7% of parents in the Current SD group, and 52% of those in the Past SD group, reported that they have felt threatened or vulnerable due to an interaction with a member of school staff (see Figure 6D). A Kruskal-Wallis H test with Mann-Whitney U post-hoc analyses revealed that significantly more parents in the Current and Past SD groups have felt threatened or vulnerable compared to parents in the No SD and Lifelong EHE groups ([Current SD/Past SD]>[No SD/Lifelong EHE]).

### 3.3. Action taken against parents to enforce attendance

When asked about action taken against parents to enforce school attendance, 42.1% of the professional group who responded to this question stated that they (or a colleague working with them on a case) had taken action against a parent/parents to enforce school attendance that had resulted in a fine (sometimes known as a ‘penalty notice’), 21.1% had taken action that resulted in a ‘Child in Need’ assessment being conducted, 10.5% reported having taken action that resulted in ‘Child Protection’ procedures, with a similar percentage having taken action that had resulted in a Parenting Order being issued, and 5.3% of the professional group reported having taken action that resulted in a School Attendance Order and a Community Order. Separately (but relatedly), 18.7% of the professional group reported having punished (or given sanctions) directly to a CYP because of their school attendance record, and 63.2% had rewarded CYP for 100% school attendance (hence indirectly penalising students with attendance difficulties).

Considering parent responses regarding specific actions taken against them because of their child’s school attendance, most of the School Distress parents in our cohort reported that ‘no action’ had been taken against them by professionals. However, a small percentage of parents reported being fined, having a School Attendance Order issued against them, being accused of Fabricated or Induced Illness (Fii), and/or facing involvement from Child Protection Services (for full details see Table 5).

**Table 5:**
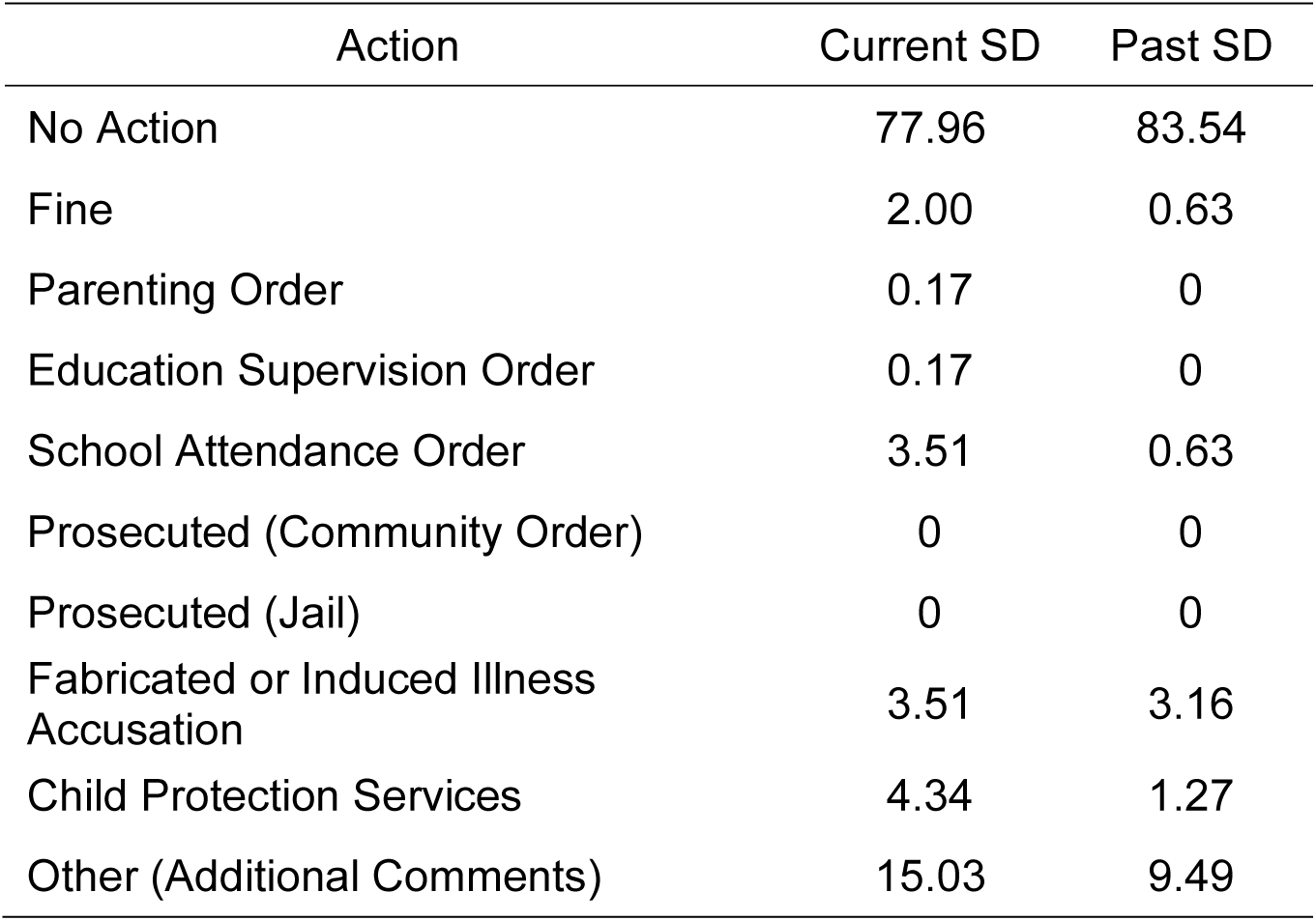
Percentage of Parents Who Have Had Different Types of Action Taken Against Them to Enforce School Attendance.

#### 3.3.1. Thematic Analysis (Parental Experiences)

Outside of the response options that we provided to parents and professionals, 106 School Distress parents provided additional comments in response to this question. Given the breadth of information provided here, a thematic analysis was conducted, with four themes (‘Dread, Fear and Vulnerability’, ‘Hostile Action(s)’, ‘Protection’, ‘Dereliction of Duty’) and eight subthemes identified. Figure 7 displays the thematic map that highlights the links between the themes and subthemes. Additional example quotes from each theme/subtheme are presented in Table 6.

**Figure 7:**
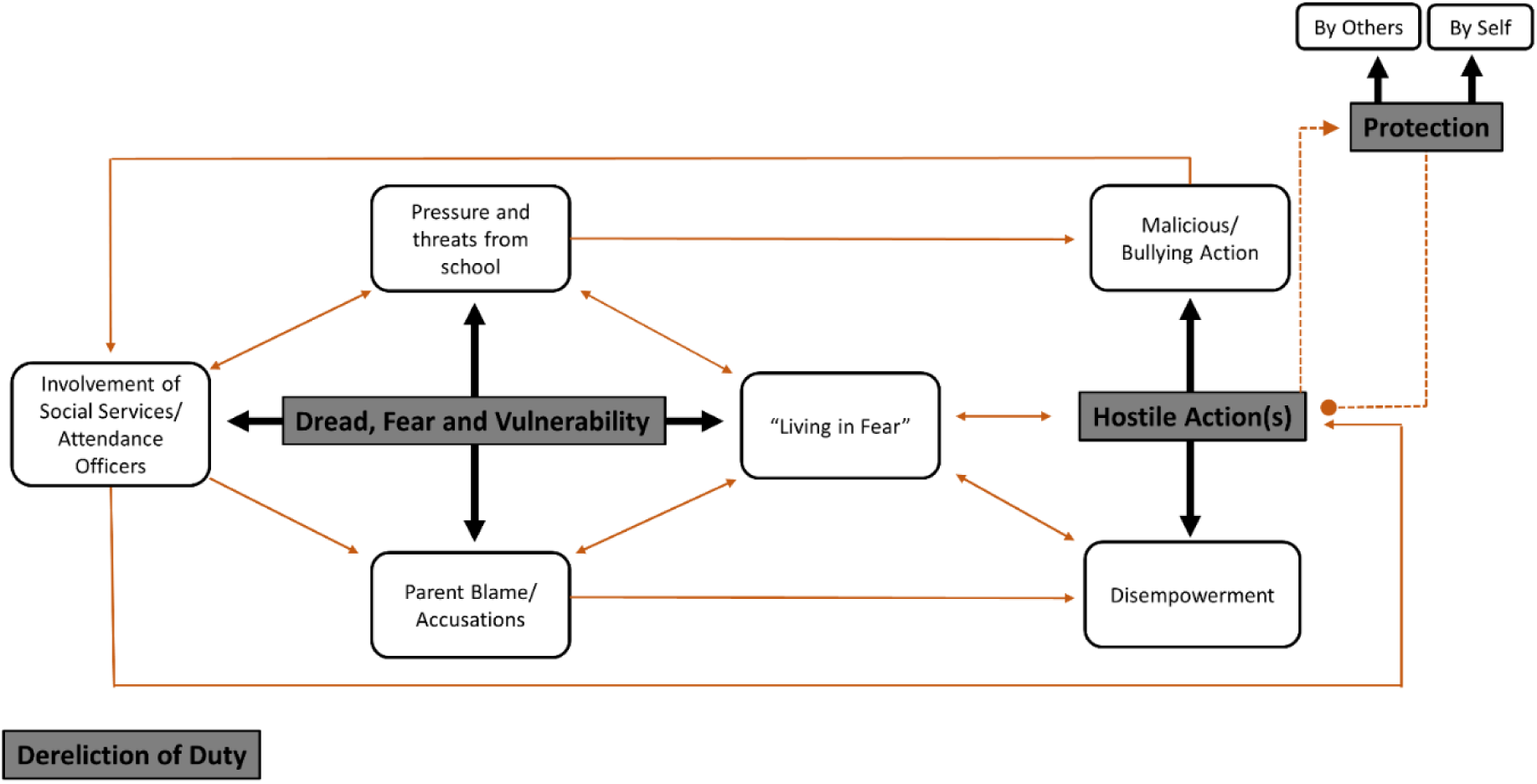
Thematic map representing the actions taken by professionals in response to School Distress. Themes are represented in the dark grey rectangles, whilst the sub-themes are represented in white boxes. The thick, black arrows indicate the direction of relationships between Themes and Sub-themes, whilst the lighter, brown arrows indicate the direction of relationships between sub-themes or from a sub-theme to themes. To arrive at the themes, each comment was read multiple times and labelled with a code. Where appropriate, comments were split apart, and each section was given a separate code. Coding focused primarily on the semantic content of comments, extracting parents’ explicit accounts, rather than any latent meanings in the data. A total of 31 codes were identified. Codes which dealt with similar issues were clustered to form initial themes and data relating to each theme was gathered. Themes were discussed and refined by the research team until consensus was reached, ensuring the themes made sense in terms of the coded extracts and the whole data set. This initially gave rise to nine themes (see (29)). However, an additional interpretative step was later performed to more deeply reflect on the significance of the previously identified patterns, their broader meanings, and the implications of these patterns. This led to this revised thematic map, involving four themes and eight subthemes.

**Table 6.**
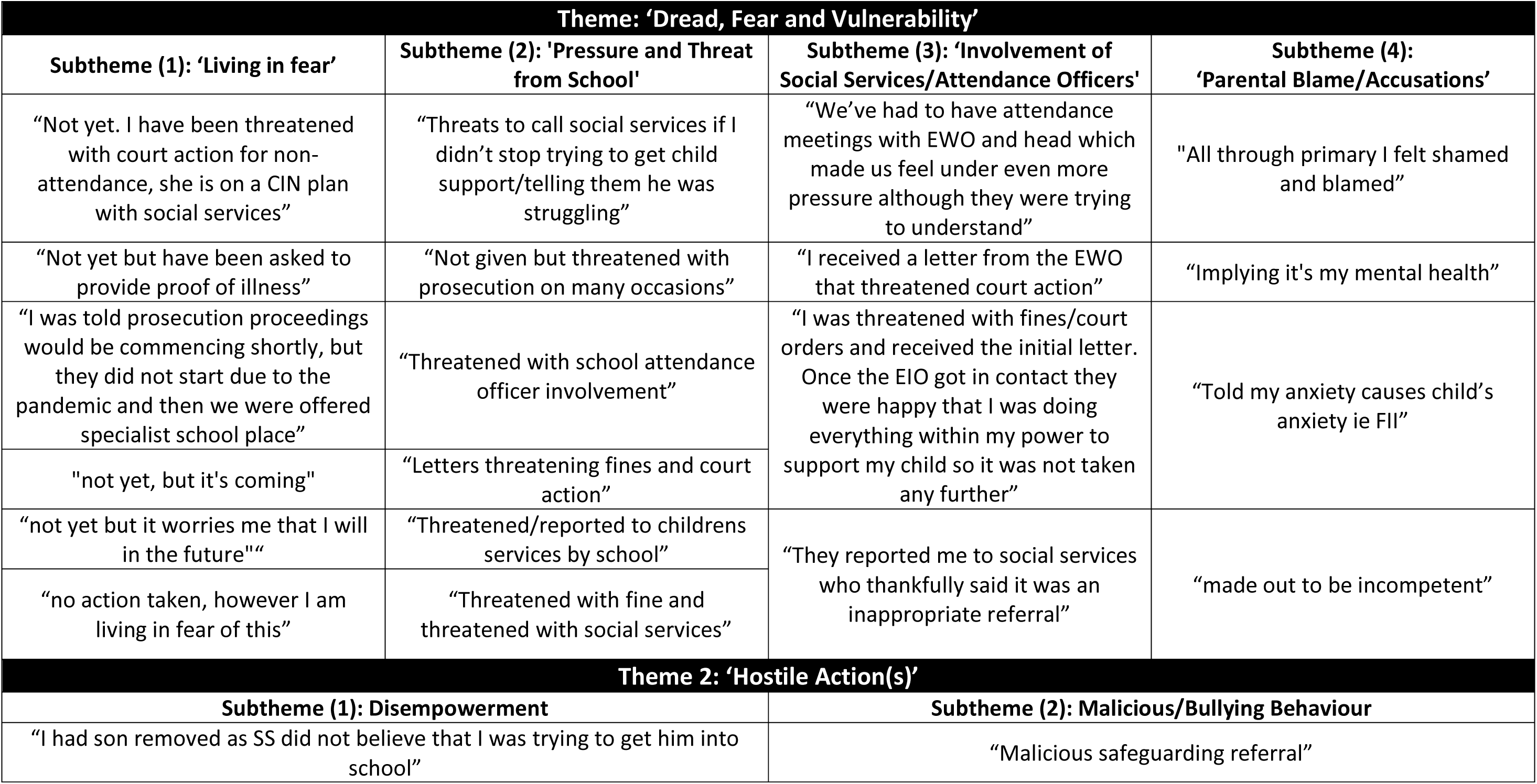
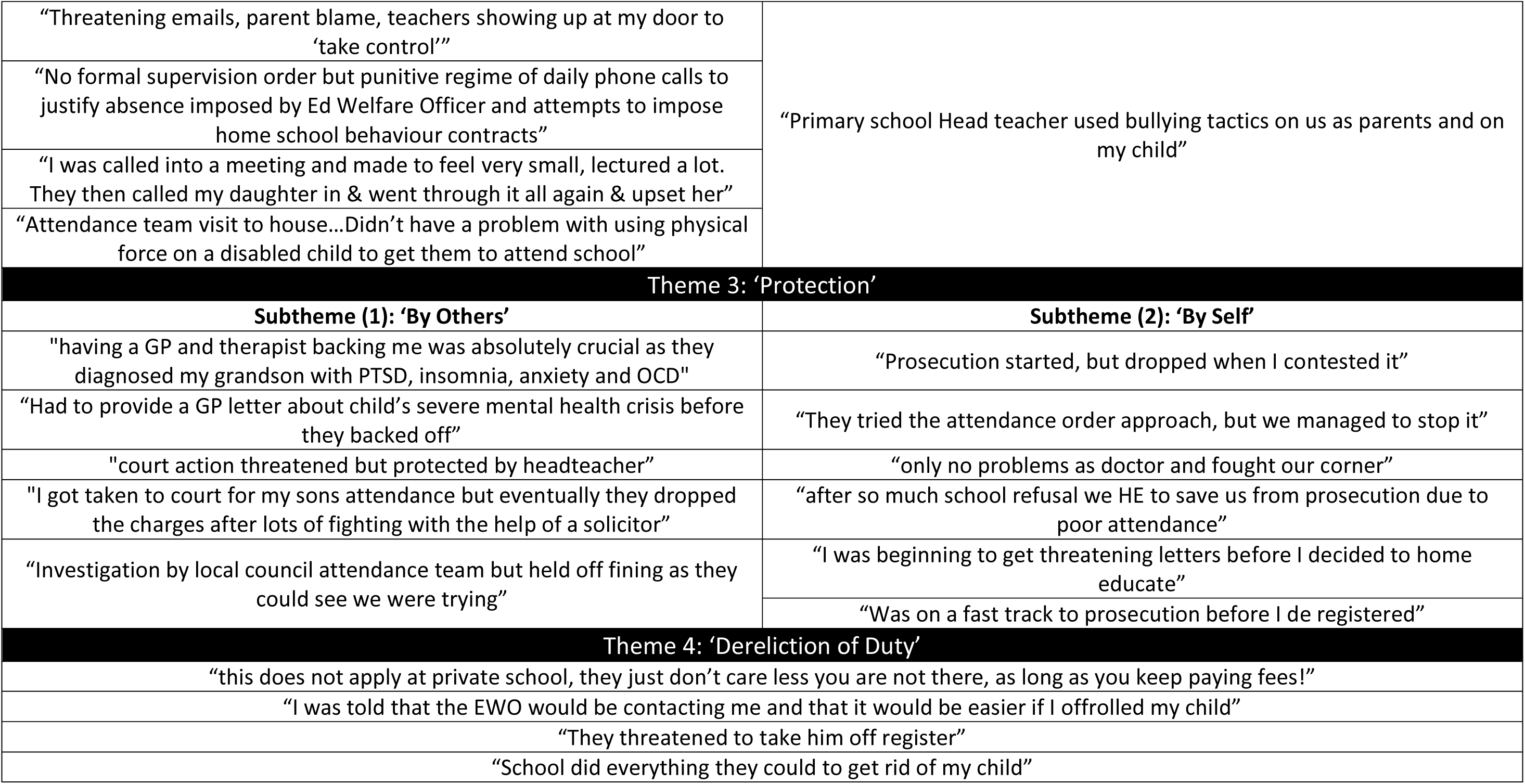
Example quotations from parent participants who have experienced School Distress, and which support the themes identified via thematic analysis. CIN = Child in Need; De register = agree to remove a child from their school setting to home educate them; EWO Education Welfare Officer; EIO = Early Intervention Officer; FII = Fabricated or Induced Illness; GP = General Practitioner/Family Doctor; HE = Home Educate; OCD = Obsessive Compulsive Disorder; PTSD = Post Traumatic Stress Disorder; SS = Children’s Social Services.

| Theme: ‘Dread, Fear and Vulnerability’ |  |  |  |
| --- | --- | --- | --- |
| Subtheme (1): ‘Living in fear’ | Subtheme (2): 'Pressure and Threat from School' | Subtheme (3): ‘Involvement of Social Services/Attendance Officers' | Subtheme (4): ‘Parental Blame/Accusations’ |
| “Not yet. I have been threatened with court action for non-attendance, she is on a CIN plan with social services” | “Threats to call social services if I didn’t stop trying to get child support/telling them he was struggling” | “We’ve had to have attendance meetings with EWO and head which made us feel under even more pressure although they were trying to understand” | "All through primary I felt shamed and blamed” |
| “Not yet but have been asked to provide proof of illness” | “Not given but threatened with prosecution on many occasions” | “I received a letter from the EWO that threatened court action” | “Implying it's my mental health” |
| “I was told prosecution proceedings would be commencing shortly, but they did not start due to the pandemic and then we were offered specialist school place” | “Threatened with school attendance officer involvement” | “I was threatened with fines/court orders and received the initial letter. Once the EIO got in contact they were happy that I was doing everything within my power to support my child so it was not taken any further” | “Told my anxiety causes child’s anxiety ie FII” |
| "not yet, but it's coming" | “Letters threatening fines and court action” |  |  |
| “not yet but it worries me that I will in the future” | “Threatened/reported to childrens services by school” | “They reported me to social services who thankfully said it was an inappropriate referral” | “made out to be incompetent” |
| “no action taken, however I am living in fear of this” | “Threatened with fine and threatened with social services” |  |  |
| Theme 2: ‘Hostile Action(s)’ |  |  |  |
| Subtheme (1): Disempowerment |  | Subtheme (2): Malicious/Bullying Behaviour |  |
| “I had son removed as SS did not believe that I was trying to get him into school” |  | “Malicious safeguarding referral” |  |

|  |  |
| --- | --- |
| “Threatening emails, parent blame, teachers showing up at my door to ‘take control’” | “Primary school Head teacher used bullying tactics on us as parents and on my child” |
| “No formal supervision order but punitive regime of daily phone calls to justify absence imposed by Ed Welfare Officer and attempts to impose home school behaviour contracts” |  |
| “I was called into a meeting and made to feel very small, lectured a lot. They then called my daughter in & went through it all again & upset her” |  |
| “Attendance team visit to house...Didn’t have a problem with using physical force on a disabled child to get them to attend school” |  |
| Theme 3: ‘Protection’ |  |
| Subtheme (1): ‘By Others’ | Subtheme (2): ‘By Self’ |
| "having a GP and therapist backing me was absolutely crucial as they diagnosed my grandson with PTSD, insomnia, anxiety and OCD" | “Prosecution started, but dropped when I contested it” |
| “Had to provide a GP letter about child’s severe mental health crisis before they backed off” | “They tried the attendance order approach, but we managed to stop it” |
| "court action threatened but protected by headteacher” | “only no problems as doctor and fought our corner” |
| "I got taken to court for my sons attendance but eventually they dropped the charges after lots of fighting with the help of a solicitor” | “after so much school refusal we HE to save us from prosecution due to poor attendance” |
| “Investigation by local council attendance team but held off fining as they could see we were trying” | “I was beginning to get threatening letters before I decided to home educate” |
|  | “Was on a fast track to prosecution before I de registered” |
| Theme 4: ‘Dereliction of Duty’ |  |
| “this does not apply at private school, they just don’t care less you are not there, as long as you keep paying fees!” |  |
| “I was told that the EWO would be contacting me and that it would be easier if I offrolled my child” |  |
| “They threatened to take him off register” |  |
| “School did everything they could to get rid of my child” |  |

The first theme, ***‘Dread, Fear and Vulnerability’*** encapsulates the feelings of dread and fear that parents feel because of threats they receive, mainly from school staff, about being fined or prosecuted for their child’s non-attendance at school or being referred to external agencies such as School Attendance Officers and/or Children’s Social Services. It also encompasses parental vulnerability; specifically, their vulnerability to being blamed for their child’s school attendance difficulties and/or accused of child abuse. Within this theme, four interlinked subthemes were identified:

- ***“Living in Fear”*** - Despite no action having been taken yet, parents often reported being concerned about punitive actions that may be taken against them in the future. The words “not yet” appeared frequently in these descriptions e.g., “*Not yet. I have been threatened with court action for non-attendance, she is on a CIN* [Child in Need] *plan with social services*”. The extract “*no action taken, however I am living in fear of this*” further underlined the dread of impending action and the fear that this inserts into parents’ daily lives.
- In ***‘Pressure and Threat from School’***, parents frequently reported being placed under pressure to enforce attendance (e.g., “*constant pressure from school*”, “*Just continuous letters from school*”). In addition, the word “*threatened*” was pervasive (i.e., it was mentioned 40 times within the 106 individual responses). These threats took the form of threats of fines, threats of prosecution, or threats of being reported to external agencies such as School Attendance Officers and/or Children’s Social Services (presumably for investigation and/or prosecution).
- A closely related subtheme was labelled ***‘Involvement of Social Services/Attendance Officers’***. This related to actual referrals of the families to external agencies by school (e.g., “*School contacted MASH* [Multi-Agency Safeguarding Hub] for Child in Need assessment”). In some instances, this led to additional threats and pressure. Other quotes indicated that although the threat of referral had been carried out by school staff, the outcome was non-threatening.
- Finally, within this theme, the subtheme ***‘Parental Blame/Accusations’*** encompassed parental vulnerability to accusations and/or blame as a result of their child’s school attendance difficulties, for example “*All through primary I felt shamed and blamed*” and “*Told my anxiety causes child’s anxiety ie FII* [Fabricated or Induced Illness]”. Such allocation of blame and/or accusations were reported to have been received from multiple sources (e.g. head teachers, family doctor).

The second theme, ***‘Hostile Action(s)’***, encompasses the instances where the situation progressed beyond threats and accusation to action against parents. Considering the level of severe emotional distress, clinically significant anxiety symptomology, and the disability-related barriers to school attendance evident within this cohort of CYP (see (5) for full description), punitive actions against their parents within this context can be perceived as hostile. Example quotes include “*I have been told I have to attend a formal interview under caution*” and “*I got taken to court for my sons attendance”.* Reflecting further on parental descriptions of actions taken, we arrived at two subthemes within this broader theme:

- The subtheme ‘***Disempowerment***’ arose as some descriptions reflected a loss of parental autonomy as a consequence of the actions taken by professionals to enforce school attendance. Example quotes include “*Threatening emails, parent blame, teachers showing up at my door to ‘take control*’”. In addition to disempowering parents, some of the actions described were likely to have caused significant trauma to the parent(s) and child, for example “*Attendance team visit to house…Didn’t have a problem with using physical force on a disabled child to get them to attend school*”.
- ‘***Malicious/Bullying Behaviour’*** arose from descriptions that were distinct from other actions described, in that the motivation for the described action taken against parents was perceived by the parents as stemming from a place of ill intent, e.g., “*Malicious safeguarding referral*”.

Running alongside the above, a third theme was identified, labelled as ‘**Protection**’. This highlighted instances where action was taken to protect parents from punitive action, disempowerment and/or prosecution. Within this, there were two distinct subthemes:

- Protective actions taken ‘***By Others’*** (i.e., by a specific professional surrounding the family). For one parent this was a member of school staff (“*court action threatened but protected by headteacher*”), whilst the others referred to external professionals, including doctors and solicitors.
- The other subtheme that emerged (labelled ‘***By Self’***) highlighted instances where parents successfully defended themselves against such actions. Example quotes included “*Prosecution started, but dropped when I contested it*”. Interestingly, in some cases, self-protection required parents to remove their child entirely from the state education system (i.e., by agreeing to home-educate their child).

Whilst the above themes and subthemes appeared interconnected, the final theme labelled ‘***Dereliction of Duty***’ appeared independent. This described instances where parents reported that their child’s school were either uninterested in the child’s lack of school attendance (e.g., “*No this does not apply at private school, they just don’t care less you are not there, as long as you keep paying fees!*”), or where schools tried to ensure that the child left the school (e.g., “*I was told that the EWO would be contacting me and that it would be easier if I offrolled my child*”, “*They threatened to take him off register*”, and “*School did everything they could to get rid of my child*”).

### 3.4. Causal Factors

Notable similarities and differences emerged when School Distress parents and professionals were asked to identify the potential causal factors of School Distress (see Figure 8), with both School Distress parent groups and the professional group ranking ‘Anxiety’ in top position. Despite this similarity, professionals were significantly more likely to select ‘parent-related’ factors than parents. More specifically, professionals selected ‘Parental mental health’ (X^2^ (1, N = 614) = 94.409, *p* < .001), ‘Over-dependency within the family’ (X^2^ (1, N = 614) = 31.367, *p* < .001), ‘Overprotective parenting’ (X^2^ (1, N = 614) = 62.836, *p* < .001), ‘Difficulties at home/within the family’ (X^2^ (1, N = 614) = 62.836, *p* < .001), and ‘Poor parenting/lack of discipline’ (X^2^ (1, N = 614) = 62.836, *p* < .001) significantly more frequently than School Distress parents (see Figure 8C and 8D). Professionals were also significantly more likely to select ‘Separation difficulties’ (X^2^ (1, N = 614) = 9.996, *p* < .01), ‘Illness’ (X^2^ (1, N = 614) = 4.804, *p* < .05), ‘Lack of Friendships’ (X^2^ (1, N = 614) = 8.456, *p* < .01), and ‘Non-Compliance’ (X^2^ (1, N = 614) = 22.897, *p* < .001) than School Distress parents.

**Figure 8:**
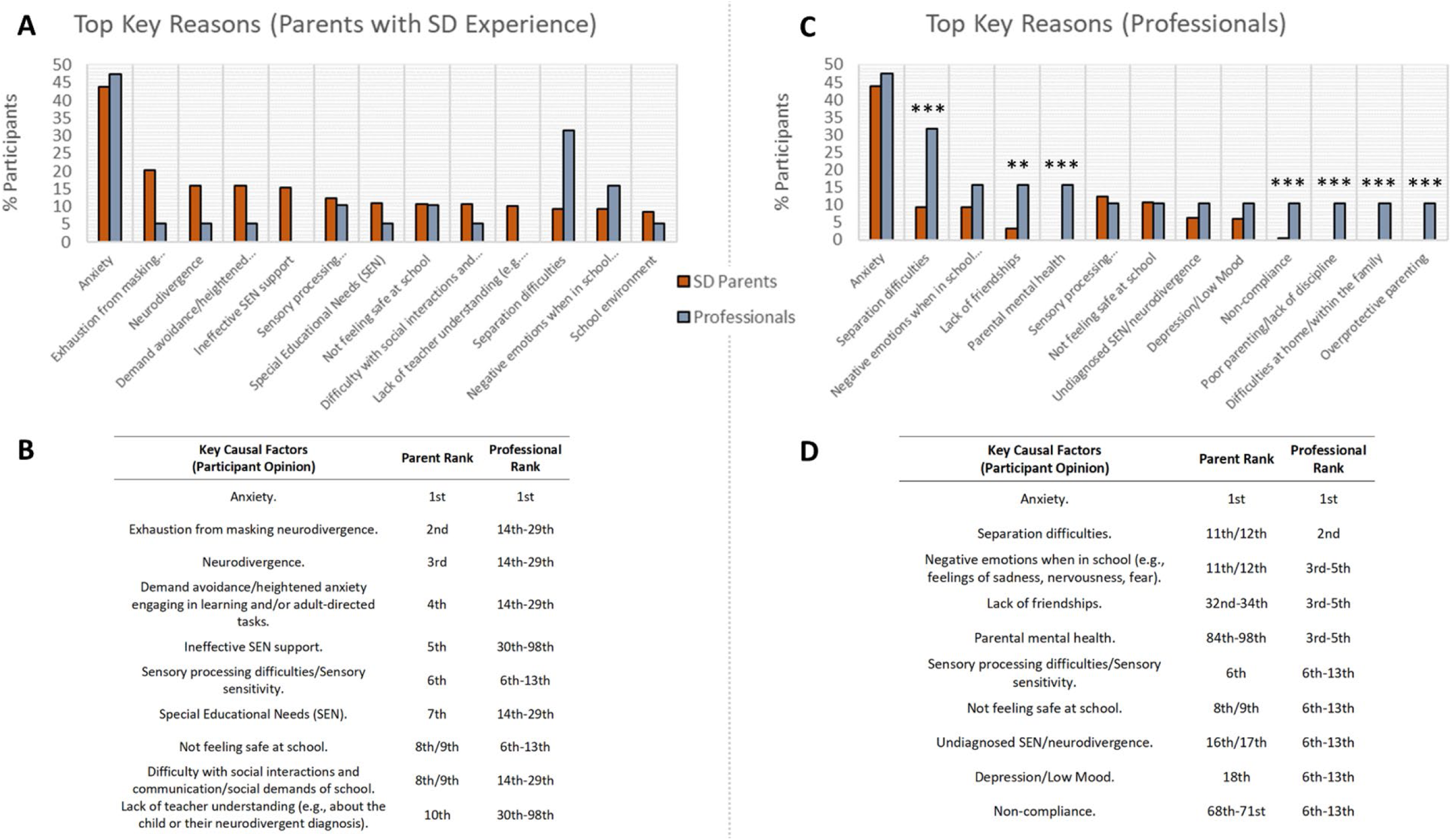
**Panels A and B**. The 13 highest ranked ‘key’ reasons identified by parents of CYP with School Distress, presented alongside the rankings provided by the professional group for the same items. **Panels C and D**. The 13 highest ranked ‘key’ reasons identified by professionals, presented alongside the rankings provided by parents with experience of SD for the same items. Significant between-group differences in how frequently these items were endorsed by these two groups are highlighted: ** p < .01, *** *p* < .001.

In contrast, School Distress parents were significantly more likely to select ‘Intolerance of uncertainty (tendency to react negatively to uncertainty)’ (X^2^ (1, N = 614) = 8.456, *p* < .01) than professionals. Moreover, after ‘Anxiety’, the most frequently identified key reasons for School Distress by School Distress parents were ‘Exhaustion from Masking Neurodivergence’ (2^nd^), ‘Neurodivergence’ (3^rd^), ‘Demand Avoidance’ (4^th^), ‘Ineffective SEN support’ (5^th^), ‘Sensory Processing Difficulties’ (6^th^), and ‘Special Educational Needs’ (7^th^) (see Figure 8A and 8B, and Supplementals Notes 4 and 5). It is noteworthy that there are all disability-related factors, revealing a pattern which differed greatly from that found in the responses of the Professional group. Finally, a ‘Lack of teacher understanding (e.g., about the child or their neurodivergent diagnosis)’ was rated 10^th^ by parents but was one of the reasons least likely to be selected by the Professional group.

### 3.5. Sources of Support

In response to the question “As a parent, what has been your most important source(s) of support?”, 39.25% of School Distress parents referred to other parents with similar experiences/parent support groups (e.g., Facebook communities), 15.96% referred to their own family/husband/partner/their CYP’s other parent, and 11.75% referred to their friends. Additional responses from a smaller number of parents included teachers, SENCOs, support workers, SENDIASS, GPs, clinical/educational/private psychologists and private therapists, as well as the internet/their own research. Concerningly, several parents also highlighted having received no support, or having to supports themselves.

Professionals were also asked to identify their most important source(s) of support and how this support could be improved (see Table 7 for professional quotations). Many professionals reflected back the same lack of support that the parents frequently referred to. They also described barriers to accessing what available support there is for CYP experiencing School Distress including lengthy referral processes, long waiting times, and external support that typically comes too late (e.g., “*they do not engage sometimes not until crisis point*”). Whilst some professionals referred to the parents/families as an important source of support, and working alongside families and external agencies as helpful, this was, however, contingent on them having the time to do this and/or access to external agencies in the first instance. CAMHS, Early Help, Pastoral staff, educational therapy, networks, and school counselling teams were all referenced.

**Table 7:** Professionals’ reflections on help and support available to them and the CYP and families they are supporting.

| Lack of Support: |  |  |  |  |  |
| --- | --- | --- | --- | --- | --- |
| “There is little to no support” | “Other than LA fines, no support from higher level. School leadership have to play good cop/bad cop” | “There is little or no support for schools in tackling attendance difficulties. Local authority intervention has been reduced even further this year... Schools are operating alone” | “It is often a box ticking exercise. Real help isn’t easy to access” | “We simply need more of them- it is worrying when parents look to me for support and provision of services and they are placed on a waiting list” |  |
| Barriers to Support: |  |  |  |  |  |
| “external services need to have a swift referral process rather than it be lengthy as professionals have limited capacity to fill in forms to acquire the help necessary” | All services need to work together and it cannot just be left to mental health services or schools by themselves to support CYP to get back in to school | “on several occasions [external services] have not picked up families and it has taken the police to force them even though trained professionals have referred in with concerns which are ignored or sign posted to agencies that are not appropriate” | “Time to work with families and children” | “Early intervention could prevent poor attendance habits from becoming established early for the CYP” | “Knowing fully what’s available and what to do of everything has been tried” |
| Positive Experiences: |  |  |  |  |  |
| “Meetings with parents/carers so we can both come up with a solution” |  |  | “Working alongside family and CAMHS” |  |  |

Almost half (46.7%) of the professional group indicated that they would like more support to help CYP with school attendance problems, whilst 60% indicated that they would like more training. A similar proportion of the professional group (60%) indicated that they would also like more training to support autistic CYP (see Table S2).

## 4. Discussion

This study shines a valuable light on the experiences of parents of CYP experiencing School Distress in the UK, including the treatment of parents by professionals, action taken against parents due to their child’s School Distress, beneficial sources of support, and the impact of the experience on the parents themselves and their wider family. Valuable input was also provided by a separate group of professionals with experience of working with CYP with school attendance difficulties, enabling a comparison of parental lived-experiences and understanding of School Distress with the lived-experiences and understanding of professionals.

Findings revealed the significant deleterious impact that School Distress can have on multiple aspects of parent’s lives, including on their mental and physical health, their careers, their financial situation, their other children, and their relationships with their partner, their family, and their friends. Strikingly, over half of parents in the Current SD group reported that they developed a new mental health condition since the onset of their child’s School Distress.

The parents’ narrative descriptions of impact focused heavily on harm to parental mental health. This was often driven by a loss of confidence in their parenting abilities and a loss of confidence in themselves, including attenuated self-esteem and self-belief. In addition, parents described the loss of leisure time, friendships (both personal friendships and their child’s friendships), sexual relationships, self-care, time to support their own mental health needs, and their ability to carry out normal daily activities. Parents also noted a negative impact on their partner’s mental health and career, and multiple parents referred to breakdowns in their relationship with their child who is experiencing School Distress. Moreover, parents often described a profound loss of trust in the system and in professionals working within this system; a loss previously documented by parents of disabled children in discussions surrounding their interactions with children’s social services e.g., “*Parents have lost trust and so have I*” (30). Here we extend this finding to include the breakdown of trust that parents have for educational systems, staff and other professionals. Only one parent noted a positive impact on their life, namely that they get to be at home more now. Analysis of responses in the professional group did not reveal any areas where professionals were significantly and deleteriously impacted as a result of working with CYP experiencing school attendance difficulties.

In addition, relative to parents whose children are not currently experiencing School Distress (i.e., parents in the No SD control group, the lifelong EHE group, and the Past SD group), parents in the Current SD group reported significantly poorer *current* mood and significantly higher *current* anxiety levels. Analyses exploring changes in mood and anxiety ratings at different points in time (i.e., Current School Distress Group: Before and During; Past School Distress Group: Before, During and After) revealed that parents’ mood declined significantly during their children’s School Distress and anxiety levels increased significantly. Whilst it is important to note that some of these ratings were provided retrospectively, and that retrospective reporting may have led to inaccuracies (31), the strikingly similar pattern observed across the Past and Current SD groups is notable, particularly given the differing temporal perspectives of these two groups of parents.

One possible factor of relevance here is the high rate of neurodivergence in the parents of CYP in both School Distress groups, as there are strong links between neurodivergence (ND) and mental health difficulties such as anxiety and depression (32–34). However, whilst parental ND may be relevant, it cannot explain the specific pattern of mood and anxiety results reported here, which deteriorated specifically during active periods of School Distress. Similarly, whilst there were comparable rates of neurodivergence in parents in the Current SD, Past SD and Lifelong EHE groups, it was only the parents in the Current SD group who demonstrated the striking elevation of *current* anxiety and the significant attenuation of *current* mood.

Poor mental health in parents of CYP with school attendance difficulties has been reported elsewhere in the literature (e.g., (35)), however such observations have been used to infer that poor parental mental health plays a causal role in the CYP’s School Distress. The temporal pattern of decline in parental wellbeing observed here challenges this inference, and instead suggests that poor parental mental health may not (at least at the group level) precede the experience of School Distress, but instead occur specifically during active periods of School Distress in their children. Hence, heightened anxiety and low mood may be a consequence of the parental experience of School Distress, as opposed to a precipitating factor of the child’s School Distress in the first instance. Future research should further explore the directionality of this effect, ideally using longitudinal assessment of parental mental health across multiple time points. Such research should also consider parental neurotype, as the significantly elevated rate of neurodivergence amongst School Distress parents may help explain the higher rates of parental mental health difficulties (36) previously noted in the School Distress literature (37).

To gain further traction over the parental lived experience of School Distress, we asked parents to rate this experience with respect to other life events containing considerable long-term contextual threat. The result of this comparison was striking, with parents currently supporting children with School Distress rating this life experience as the second most threatening life event, superseded only by the experience of a ‘Death of a 1st Degree Relative, including spouse or child’, and more threatening than events such as ‘Death of a close family friend’, ‘Serious Illness or Injury to Self’, and ‘Serious Illness or Injury to Close Relative’. Both School Distress parent groups and the professional group rated the experience of a child “school refusing” as significantly more threatening than parents in the two control groups who had no direct lived experience of School Distress (i.e., the No SD and Lifelong EHE groups). This mismatch is in keeping with the wider lack of societal understanding of School Distress.

Reinforcing this level of threat was the finding that parents supporting CYP with School Distress had significantly higher levels of all negative emotion states measured by the Discrete Emotions Questionnaire (24) (i.e., fear, sadness, anxiety, anger, and disgust) and significantly lower levels of positive emotion states (i.e., relaxation and happiness), relative to control parents and professionals. The fear and vulnerability of parents supporting CYP with School Distress was further evidenced by the fact that parents in both the Current SD and Past SD groups reported feeling threatened or vulnerable due to an interaction with a member of school staff significantly more frequently than control parents, and is further delineated in the thematic analysis discussed below.

The two School Distress parent groups were also significantly more likely to report not being believed by school staff, health professionals and other professionals (including Local Authority staff and Children’s Social Services) when raising concerns about their child’s difficulties, and to experience professional gaslighting, relative to parents without experience of School Distress. This was supported by free text comments such as “*Dismissed and told they are fine in school*”, *“…they minimise my concerns*” and “*As my child masks in school I often get a look from the teachers and told he doesn’t do that in school he’s playing you up*”. These quotes echo the parent voices documented by Bodycote (13), who reported a dismissal of their concerns about the impact of the school environment on their children by school staff, who instead tended to blame the child’s difficulties within the school environment on deficient parenting abilities and/or problems at home, and the Hackney cohort of parents who overwhelmingly reported feeling judged, ignored, and blamed by school staff for their child’s school attendance difficulties (8).

Concerningly, over a quarter (28.6%) of participants in the professional group also reported not being believed by school staff when raising concerns about a CYP’s difficulties. Research is needed to better understand why some teachers appear reluctant to accept parent or professional opinions with respect to potential difficulties being experienced by their pupils. The pervasive view that School Distress stems from parental, family and/or child behavioural issues (14) is likely highly relevant here, and highlights the urgency of disseminating research into the complex drivers of School Distress (which often include child disability-related factors (5)) amongst school staff, especially when one considers that access to supports/interventions for children and families experiencing School Distress is typically gatekept by professionals (38).

This pervasive belief that School Distress stems from parental issues was further evidenced by the finding that the professional group in this study was significantly more likely to select ‘parent-related’ factors such as ‘parental mental health’, ‘over-dependency within the family’, ‘over-protective parenting’, ‘difficulties at home/within the family’, and ‘poor parenting/lack of discipline’ to be drivers of a CYP’s School Distress, relative to the School Distress parents, with one participant in the professional group stating:

> *“We are seeing more attendance difficulties with students in year 7, 8 and 9 students particularly. Parents/carers with their own anxieties tend project onto their children. I feel that even after every reasonable adjustment it is never enough for these parents”* (SENCO).

Moreover, whilst ‘overprotective parenting’, ‘poor parenting/lack of discipline’, and ‘to gain attention from a parent/caregiver’ emerged amongst the most frequently identified causal factors by the professional group, these reasons were amongst the least frequently selected by parents. This finding corroborates previous research in which parents have described feeling unjustly blamed for their children’s school attendance difficulties (8, 13, 16–18). However, it is important to note that one participant in our professional group (a senior mental health lead for a service working with young people who are experiencing School Distress and are unable to attend school) actively advocated against professionals orientating towards parental blame explanations, stating: *“The parents are under enormous stress-we must avoid blaming parents*”.

Professionals were also significantly more likely to highlight child behaviour factors such as ‘non-compliance’ than parents, situating the problem within the child themselves (14). This, again, is consistent with previous parental and child reports (17, 18). Parents, on the other hand, were significantly more likely than professionals to identify the disability-related factor ‘intolerance of uncertainty’ as a factor underlying School Distress. Intolerance of uncertainty is a transdiagnostic risk and maintaining factor for emotional disorders (39), is a central feature in anxiety-related experience (40), and has been proposed as a framework for understanding anxiety in autistic children and adolescents (41) and therefore it is vital that this is not ignored. The finding that disability-related factors were more strongly endorsed by parents, relative to professionals, is consistent with previous studies conducted with parents and young people themselves, with their findings highlighting factors such as anxiety, trauma, social exclusion/isolation, the sensory environment of school, and fear of teacher behaviour as causing or contributing to School Distress (42–46).

Poor understanding of autism can also explain why ‘separation difficulties’ was selected significantly more frequently by professionals than by parents, as ‘separation difficulties’ in neurotypical children often occur as a consequence of atypical early life attachment, whereas in autistic children, they can relate to sensory hyperactivity (47, 48). As teachers and other professionals often have a good educational grounding in attachment theory in neurotypical children, but not of autism (49, 50), they are thus primed by their training to misattribute autistic children’s distress at being separated from their safe adult at the school gate to attachment difficulties with their care-giver, and hence more likely to generate parental blame explanations of such presentations. Indeed, the majority of participants in our professional group indicated that they personally would like more training to support autistic CYP.

The tone of conversations between parents of children with School Distress and school staff also warrants further research investigation and likely the provision of additional training for school staff as this was described using strikingly different terms (e.g., ‘dismissive’, ‘critical’, ‘unsupportive’, ’uninformed’, ‘patronising’, ‘deceitful’, ‘condescending’ …etc.) by parents of CYP with School Distress, relative to control parents (who described these conversations as ‘friendly’, ‘calm’, ’caring’, and ’helpful’). The fact that it is overwhelmingly parents of disabled children who are reporting these negative tones from their children’s school staff is of extra concern, and aligns with previous research that reported that when seeking an initial educational placement for their child, parents of autistic children often feel intimidated by school staff and not believed regarding their child’s difficulties (51).

Of potential relevance too is the finding that participants in our School Distress parent groups were significantly more likely to be neurodivergent than control parents. For neurotypical teachers, interactions with autistic parents are likely experienced as unusual (52) and rapidly perceived as less favourable than interactions with neurotypical parents (53). These cross-neurotype differences in communication styles may appear to professionals working with parents of children experiencing School Distress as markers of “family dysfunction” (54) when perceived through a neurotypical lens, hence leading to more negative responses from professionals. Research exploring relationships between parent-teacher pairs that share the same neurotype versus those that have different neurotypes would likely be helpful here.

Potentially relevant too are the findings of a comparative study of autistic and non-autistic women’s experiences of motherhood (36). This study found that autistic mothers were also more likely to report feeling misunderstood by professionals and reported experiencing higher levels of anxiety and selective mutism when interacting with professionals. They also had concerns with respect to not knowing which details were appropriate to share with professionals. Increased recognition and understanding of parental neurodivergence, including the strong bonds and intense connection and love that autistic mother’s report sharing with their children (55), may also help prevent unnecessary safe-guarding referrals of School Distress families into children’s social services.

The fear and vulnerability of parents supporting CYP with School Distress was also evident in our thematic analysis within the core theme of ‘Dread, Fear and Vulnerability’. Parental awareness of the power that professionals have to initiate punitive actions against parents of children with poor school attendance (which can range from fines to imprisonment) often drove this dread, fear and vulnerability, as did the threat of having one’s parenting capacity questioned by others, including children’s social services. Similar findings have been reported in research on children’s social services, for example, “*there was a tendency to use the social work assessment as an opportunity to judge parenting capacity through a child protection lens rather than through a lens of social care need. This has long been a complaint of families caring for disabled children*” (56) (pg. 13; see also (30)).

The related theme of ‘Hostile Action(s)’ and subtheme ‘Malicious/Bullying Behaviour’ extended the above, with some parents describing the use of what they perceived to be malicious practice by school staff. Again, such behaviour has been reported elsewhere e.g., “*a growing number* [of parents] *are finding that they are being investigated for fabricated and induced illness as a result of challenging the practice*” (30), requesting support for their disabled child, or raising a complaint about a lack of support (57).

Within this second theme of ‘Hostile Action(s)’ was the subtheme ‘Disempowerment’. This encompassed scenarios where parents described a loss of parental autonomy because of the actions taken against them by professionals to enforce school attendance. This may help explain the high levels of new mental health conditions (1 in every 2 parents) reported by parents in the Current School Distress group, as disempowerment is an essential feature of psychological trauma (58–60).

Moreover, although there were no quotes linking the decision to de-register a child from school into Home Education directly with the sub-theme of parental disempowerment, many quotes indicated that there was a connection between the themes of ‘Hostile action(s)’ and ‘Dread, Fear and Vulnerability’ and parents deciding to remove their children from their school roll to home educate them. Future research is urgently required to establish whether the threat of punitive action(s), disempowerment and or malicious/bullying behaviour is knowingly being used by educational professionals to ‘off-roll’ CYP perceived as ‘problematic’ from the school, and if so, to establish how widespread this practice is. Off-rolling (i.e., excluding children from school for non-disciplinary reasons) is unlawful in the UK, although respective UK governments are aware that this unlawful practice does occur (61–63). Alongside compelling evidence of increasing School Distress amongst UK school children, such links may explain why there is a simultaneous increasing rate of home-education in the UK (64, 65). Recent data collected by the home education charity ‘Education Otherwise’ is revealing in this regard – with over half (54%) of parents who had begun home education in 2023 reporting ‘schools not meeting their child’s needs’ as their primary reason for deciding to home educate their child (66), an increase from just under one-third of parents (32%) who cited this as their primary reason previously (67). Our previous finding, that School Distress is predominately driven by complex neurodivergence and unmet needs in school settings (5), alongside the almost doubling of home educating families in some areas since the Covid-19 pandemic (65), combine to create a very concerning picture.

Not all families have the option to home educate their child when their child’s needs are not being met at school, as this incurs a significant cost due to the necessary re-adjustment to family life and to parental careers; a cost which would be prohibitive for many families. Other families may simply feel unable to do this due to parental health, disability, caring, or educational constraints. In such circumstances, parents may experience an even greater sense of disempowerment due to limited options to alleviate their and their child’s distress, and hence may be at even greater risk of experiencing mental and/or physical health difficulties than parents who are in the more privileged position of being able to “choose” to home educate.

Whilst it was reassuring that a third theme ‘Protection’ was identified in the above thematic analysis, with some parents reporting being protected against such punitive actions by others (such as Head Teachers and GPs), often protection was reported within a context of the parent having to protect themselves. Sometimes this was possible due to societal and/or educational privilege (e.g., a respected professional occupation). Hence again, it is likely that it is the most vulnerable parents who are most at risk of the most deleterious impact of supporting a child through the experience of School Distress.

Returning to the findings that parents of CYP who have experienced School Distress reported feeling dismissed, not believed and gaslighted by school staff significantly more frequently than parents of CYP without School Distress, it is important to consider the consequences of these specific experiences on parental mental health. As the cohort reported here consisted primarily of mothers (97%), the wider literature on the impact of professional gaslighting on women is particularly relevant. A recent systematic review evaluating the experiences of medical gaslighting in women, which in this paper was defined as “*the dismissive, invalidating, and biased experiences of people with the healthcare system*”, found that such experiences lead to feelings of frustration, distress, isolation, extreme anxiety, and trauma, often resulting in patients turning away from their doctors and seeking support from online communities instead (68). Similar consequences of sustained gaslighting of women in other contexts have been reported elsewhere (e.g., (69)). Our findings echo these consequences, with online support groups being the most consistent source of support identified by School Distress parents. However, our parents did not just report being dismissed and invalidated by school staff, they also reported not being believed by family, friends, local authorities, social workers, psychologists, partners, ex-partners, other parents, work colleagues, and even by their own parents. In the words of one parent, they were not believed about their child’s difficulties by “*just about everyone*”. Alongside disempowerment, the other essential feature of psychological trauma is isolation (58–60).

### Strengths, Limitations, and Future Research

One limitation of the present study is that most participants were mothers, meaning that our findings regarding the parental experience of School Distress may not be representative of the experience of fathers. Future research should therefore explore the experiences of fathers, as any differences may have implications for the support offered to parents of children experiencing School Distress (70). However, it may also be telling that most respondents were mothers, as some mothers commented on how differently their partner was treated by professionals, and others called for further research on the topic to explore whether the treatment that mothers experience in this context may be underpinned by systemic misogyny. This study was not designed to explore these issues, however. In addition, this study was limited to the United Kingdom, further reducing generalisability of findings. Given that education systems vary internationally, the experiences of parents may differ between countries, providing an additional avenue for future research.

Finally, two of our participants in the Professionals group were also parents of children who have experienced School Distress. Hence, a minority of the participants in this group likely share similar lived experiences with the parents in the Past SD group.

Despite these limitations, this study also had several strengths, including its large sample size. This was much greater than in previous School Distress research, enabling stronger conclusions to be made. Additionally, the inclusion of the control parent and professional groups enabled important comparisons to be made.

## 5. Conclusions

To our knowledge, this is the first study of this scale to explore the familial experience of School Distress. Findings revealed that supporting a child experiencing School Distress is an overwhelmingly negative experience for parents. Deleterious impacts were evident across all aspects of parents’ lives, including on their mental and physical health, their careers, their financial situation, and their wider family, including their other children. Parental blame was also found to be rife, with hostile and punitive treatment by professionals surrounding the family compounding this experience and leading to parental disempowerment. A profound loss of trust in school staff, and in systems more broadly, was common. The responses in this study also revealed that those experiencing School Distress from the perspective of a parent perceive this experience to be one of the most threatening possible life events, surpassing even a serious illness or injury to themselves. Concerningly, one in every two parents currently supporting a child with School Distress reported developing a new mental health difficulty since the onset of their child’s difficulties.

Despite recognising the threatening nature of this experience for parents, professionals were significantly more likely to select parent/family-related factors as drivers of children’s school attendance difficulties and rated many disability-related factors as less relevant than parents of children who have experience of School Distress; thus, confirming parental reports of feeling blamed and shamed. This likely relates to the dominant narrative in the older School Refusal literature that typically overlooked child neurodivergence as a key causal factor of school attendance difficulties, alongside an under-appreciation of the high rates of neurodivergence amongst the parents of CYP with School Distress. Recognition of the most common antecedent of School Distress (i.e., unmet need at school often stemming from complex neurodevelopmental profiles (5)), alongside recognition of the daily stressors and serious threats facing the parents of CYP experiencing School Distress, is urgently required by educational, health and social care professionals, so that supportive and non-threatening relationships can be fostered with parents.

## Supporting information

see Supplementary Notes 2

## Data Availability

All data produced in the present study are available upon reasonable request to the authors

