## Supplementary material for "“I felt shamed and blamed”: An exploration of the parental lived experience of School Distress": see Supplementary Notes 2

### Supplemental Material

### Supplementary Note 1:

***Roles of professional participants included*** teachers, teaching assistants, higher level teaching assistants, SENCOs, headteachers, deputy headteachers, school nurses, attendance inclusion officers, welfare managers, child psychologists/psychotherapists, and educational psychologists.

**Table S1:** Percentage of participants in the Professional group who had experience working in the below educational settings or working with CYP accessing education outside of a school setting or CYP without access to education.

| **Professional experiences working in:** | |
|---|---|
| Mainstream schools/colleges | 100% |
| Special unit(s) within a mainstream school | 20% |
| Specialist school(s) | 20% |
| Alternative education establishments (pupil referral units or similar) | 46.7% |
| **Professional experiences working with CYP:** | |
| Accessing education support for medical absence/medical home tuition and education otherwise than at school | 33.3% |
| Who are without education due to being unable to access their education setting | 53.3% |
| Who are without education due to having a failed placement with no alternative identified | 33.3% |
| Who are without education due to waiting for a place to become available at a specialist provision | 20% |
| Who are now electively home educated due to difficulties experienced in their school setting and/or off-rolling | 20% |

**Table S2**: Professionals ratings with respect to their confidence in their ability to support CYP with school attendance problems and with Autistic CYP. Professionals were also asked if they would like more training and/or more supporting to better support these students.

| Professional Ratings | Very Confident | Confident | Not Very Confident | More Training | More Support |
|---|---|---|---|---|---|
| When supporting CYP with School Attendance Problems | 13.3% | 66.7% | 20% | 60% | 46.7% |
| When supporting Autistic CYP | 15.4% | 46.2% | 38.5% | 60% | 26.7% |

### Supplementary Note 2: Threatening Life Events (Brugha et al., 1985)

**Commonly reported prescribed events with a high proportion rated as having moderate or marked long-term threat:**

- Death of a 1st degree relative including child or spouse
- Major financial crisis
- Separation due to marital difficulties
- Broke off a steady relationship
- Sacked from job
- Problems with police/court appearance
- Serious Illness or Injury to close relative
- Something valuable lost or stolen
- Serious Illness or Injury to Self
- Death of close family friend or 2nd degree relative
- Serious problem with close friend, neighbour or relative
- Unemployed seeking work for more than 1 month

**Frequently reported prescribed events with a high proportion rated with mild or no long-term threat:**

- Minor illness or injury to self
- Had moderate financial difficulties
- Had a baby
- Started in a completely different type of job
- Moved house within own town/city

### Supplementary Note 3

#### The Extent to Which School Distress Has a Wider Negative Impact:

**(1) Negative impact on parents' physical health:**

Current SD: $t(526) = 49.15$, $p < .001$

Past SD: $t(127) = 19.22$, $p < .001$

**(2) Negative impact on parents’ relationships with their partners:**

Current SD: $t(460) = 49.01$, $p < .001$

Past SD: $t(119) = 18.28$, $p < .001$

**(3) Negative impact on parents’ careers:**

Current SD: $t(509) = 67.94$, $p < .001$

Past SD: $t(121) = 21.67$, $p < .001$

**(4) Negative impact on parents’ financial situation:**

Current SD: $t(476) = 48.58$, $p < .001$

Past SD: $t(121) = 18.43$, $p < .001$

**(5) Negative impact on parents’ other children:**

Current SD: $t(466) = 66.66$, $p < .001$

Past SD: $t(102) = 20.55$, $p < .001$

**(6) Negative impact on parents’ wider family unit:**

Current SD: $t(476) = 42.34$, $p < .001$

Past SD: $t(114) = 14.66$, $p < .001$

**(7) Negative impact on parents’ their family friends:**

Current SD: $t(413) = 29.87$, $p < .001$

Past SD: $t(97) = 11.66$, $p < .001$

**(8) Negative ‘Other’ impact(s):**

Current SD: $t(97) = 12.26$, $p < .001$

Past SD: $t(18) = 3.85$, $p < .01$

### Supplementary Note 4

Participants selected all potential causal factors that they believed underpinned their child's School Distress (parents)/children's School Distress more generally (professionals). Results presented below (1st = most frequently selected... 98th = least frequently selected).

| **Options Presented to participants** (*grouped into categories for clarity*) | **Parent's Rank** | **Professional's Rank** |
|---|---|---|
| **Mental Health:** | | |
| Anxiety | 1st | 1st |
| Separation difficulties | 43rd-45th | 2nd |
| Intolerance of uncertainty (tendency to react negatively to uncertainty) | 25th-26th | 37th-52nd |
| Depression/Low Mood | 46th | 37th-52nd |
| School-related trauma | 37th | 53rd-68th |
| Non-school related trauma | 77th | 28th-36th |
| Other mental health condition(s) | 61st-62nd | 19th-27th |
| **Physical Health:** | | |
| Physical health reasons | 75th | 89th-98th |
| Illness | 81st | 53rd-68th |
| Exhaustion | 33rd | 28th-36th |
| **Worries/Negative Emotions:** | | |
| Negative emotions when in school (e.g., feelings of sadness, nervousness, fear) | 4th | 9th-11th |
| Perfectionism/worries about poor performance | 15th-16th | 28th-36th |
| Afraid of getting into trouble | 24th | 53rd-68th |
| Afraid of something at school, e.g., fire alarm | 58th-59th | 53rd-68th |
| Feelings of embarrassment in front of others at school | 21st | 19th-27th |
| Low self-esteem | 34th | 5th |
| Stress | 9th | 9th-11th |

| | | |
|---|---|---|
| Not being listened to | 31st | 84th-88th |
| Stress relating to homework | 49th-50th | 53rd-68th |
| Assessment stress | 48th | 69th-83rd |
| Concerns re Covid-19 (e.g., becoming sick, transmitting the virus to others...etc.) | 70th | 37th-52nd |
| **School-Related Factors:** | | |
| School environment | 3rd | 7th-8th |
| Class size | 18th-20th | 37th-52nd |
| Difficulties with food/smells/eating at school | 38th-39th | 53rd-68th |
| Not feeling safe at school | 15th-16th | 37th-52nd |
| Staff:Student ratio | 47th | 89th-98th |
| Classroom disruption | 54th | 69th-83rd |
| Difficulties with unstructured periods (e.g., yard time) | 41st | 6th |
| Anxiety related to specific aspects of school (e.g., specific classes) | 30th | 12th-18th |
| Length of school day | 11th-12th | 69th-83rd |
| Attendance pressure | 51st | 69th-83rd |
| Changes to routine or staffing | 28th | 37th-52nd |
| Structure of the school day | 57th | 69th-83rd |
| Lack of structure to the school day | 82nd | 69th-83rd |
| Transition (within the school day) | 35th | 53rd-68th |
| Transition (from one year group to another) | 53rd | 37th-52nd |
| Transition (from one school to another) | 63rd | 19th-27th |
| Covid-19 related changes to school day/environment | 64th | 37th-52nd |
| Journey to school e.g., school bus | 68th | 37th-52nd |
| Excessive homework | 73rd | 84th-88th |
| Physical demands of school | 38th-39th | 53rd-68th |
| Problems with school regulations | 66th | 53rd-68th |
| An increasingly standardised education system | 18th-20th | 53rd-68th |
| **Academic Factors:** | | |
| Academic pressure | 32nd | 19th-27th |

| | | |
|---|---|---|
| Subject specific difficulties | 67th | 89th-98th |
| Difficulty accessing the curriculum | 52nd | 37th-52nd |
| **Disability-Related Factors:** | | |
| Special Educational Needs (SEN) | 10th | 12th-18th |
| Ineffective SEN support | 18th-20th | 28th-36th |
| Attention and learning difficulties | 36th | 28th-36th |
| Neurodivergence | 2nd | 12th-18th |
| Sensory processing difficulties/Sensory sensitivity | 5th-6th | 19th-27th |
| Undiagnosed SEN/neurodivergence | 43rd-45th | 28th-36th |
| Demand avoidance/heightened anxiety engaging in learning and/or adult-directed tasks | 13th | 3rd-4th |
| Difficulty with social interactions and communication/social demands of school | 5th-6th | 12th-18th |
| Exhaustion from masking neurodivergence | 7th | 53rd-68th |
| Burnout (relating to neurodivergence) | 17th | 69th-83rd |
| Being punished for behaviour relating to SEN/neurodivergence | 43rd-45th | 69th-83rd |
| Pressure to reduce/stop repetitive self-stimulatory behaviours (i.e., stimming) | 58th-59th | 69th-83rd |
| Not feeling their difficulties are believed | 25th-26th | 53rd-68th |
| School not accommodating individual needs | 22nd | 37th-52nd |
| Cognitive rigidity | 55th | 69th-83rd |
| **Peer Relations:** | | |
| Difficulties with friends/peers | 29th | 12th-18th |
| Unpredictable behaviour of peers/classmates | 14th | 69th-83rd |
| Lack of friendships | 40th | 12th-18th |
| Bullying (in-person) | 60th | 37th-52nd |
| Bullying (cyber) | 83rd | 37th-52nd |
| Peer violence | 80th | 84th-88th |
| Sexual harassment/violence | 93rd-96th | 84th-88th |
| Peer/gang intimidation and/or exploitation at school | 85th | 53rd-68th |
| Gang violence at school | 90th | 84th-88th |

| | | |
|---|---|---|
| **Pupil Behaviour:** | | |
| Non-compliance | 69th | 19th-27th |
| Conduct disorder | 88th | 69th-83rd |
| Repeated sanctions (related to behaviour difficulties) | 65th | 37th-52nd |
| **Teacher Related Factors:** | | |
| Lack of trust in teachers | 27th | 69th-83rd |
| Difficulties with staff | 49th-50th | 19th-27th |
| Teacher behaviour, e.g., shouting | 23rd | 89th-98th |
| Lack of teacher training in SEN/neurodiversity | 11th-12th | 53rd-68th |
| Inappropriate teacher training in SEN/neurodiversity | 42nd | 69th-83rd |
| Lack of teacher understanding (e.g., about the child or their neurodivergent diagnosis) | 8th | 19th-27th |
| **Reward/Punishment by School Staff:** | | |
| Use of behaviour modification strategies | 56th | 53rd-68th |
| Repeated sanctions (related to attendance difficulties) | 78th-79th | 69th-83rd |
| Use of restraint at school | 74th | 89th-98th |
| Use of seclusion at school (i.e., forced to spend time alone against their will) | 71st | 89th-98th |
| **Parent/Family Related Factors:** | | |
| Poor parenting/lack of discipline | 93rd-96th | 7th-8th |
| Overprotective parenting | 91st-92nd | 12th-18th |
| To gain attention from a parent / caregiver / other | 86th | 19th-27th |
| Difficulties at home/within the family | 87th | 9th-11th |
| Parental mental health | 84th | 3rd-4th |
| Detachment with little interaction among family members | 97th | 89th-98th |
| Isolation with little interaction outside the family unit | 78th-79th | 37th-52nd |
| Over-dependency within the family | 91st-92nd | 28th-36th |
| Child neglect | 98th | 28th-36th |

| | | |
|---|---|---|
| **Other:** | | |
| Poverty | 93rd-96th | 37th-52nd |
| Pursuit of tangible rewards outside of school | 93rd-96th | 89th-98th |
| Preference for doing activities outside of school | 72nd | 53rd-68th |
| Athletic competition | 89th | 89th-98th |
| Inability to engage in preferred routine | 61st-62nd | 28th-36th |
| Other(s) | 76th | 89th-98th |

### Supplementary Note 5

>> Next participants were asked to select the key causal factors from all factors they had initially above. Participants were advised to select no more than 3 key causal factors. Results below (1st = most frequently selected... 98th = least frequently selected):

| **Options Presented to participants** (*grouped into categories for clarity*) | **Parent's Rank** | **Professional's Rank** | **Sig.** |
|---|---|---|---|
| **Mental Health:** | | | |
| Anxiety | 1st | 1st | *p* = .762 |
| Separation difficulties | 11th/12th | 2nd | *p* = .002** |
| Intolerance of uncertainty (tendency to react negatively to uncertainty) | 60th-67th | 14th-29th | *p* = .028* |
| Depression/Low Mood | 18th | 6th-13th | *p* = .765 |
| School-related trauma | 16th/17th | 30th-98th | *p* = .255 |
| Non-school related trauma | 49th-52nd | 30th-98th | *p* = .634 |
| Other mental health condition(s) | 46th/47th | 30th-98th | *p* = .589 |
| **Physical Health:** | | | |
| Physical health reasons (Long-term) | 41st/42nd | 30th-98th | *p* = .532 |
| Illness | 61st-67th | 14th-29th | *p* = .028* |
| Exhaustion | 35th | 14th-29th | *p* = .579 |
| **Worries/Negative Emotions:** | | | |
| Negative emotions when in school (e.g., feelings of sadness, nervousness, fear) | 11th/12th | 3rd-5th | *p* = .353 |
| Perfectionism/worries about poor performance | 26th | 30th-98th | *p* = .372 |
| Afraid of getting into trouble | 43rd-45th | 30th-98th | *p* = .569 |
| Afraid of something at school, e.g., fire alarm | 61st-67th | 30th-98th | *p* = .720 |
| Feelings of embarrassment in front of others at school | 32nd-34th | 30th-98th | *p* = .429 |
| Low self-esteem | 21st/22nd | 14th-29th | *p* = .910 |
| Stress | 49th-52nd | 30th-98th | *p* = .634 |
| Not being listened to | 38th | 30th-98th | *p* = .499 |
| Stress relating to homework | 61st-67th | 30th-98th | *p* = .720 |

| | | | |
|---|---|---|---|
| Assessment stress | 53rd-54th | 30th-98th | $p$ = .660 |
| Concerns re Covid-19 (e.g. becoming sick, transmitting the virus to others...etc.) | 61st-67th | 30th-98th | $p$ = .720 |
| **School-Related Factors:** | | | |
| School environment | 13th | 14th-29th | $p$ = .625 |
| Class size | 43rd-45th | 30th-98th | $p$ = .596 |
| Difficulties with food/smells/eating at school | 55th-59th | 30th-98th | $p$ = .688 |
| Not feeling safe at school | 8th/9th | 6th-13th | $p$ = .993 |
| Staff:Student ratio | 61st-67th | 30th-98th | $p$ = .720 |
| Classroom disruption | 61st-67th | 30th-98th | $p$ = .720 |
| Difficulties with unstructured periods (e.g. yard time) | 39th/40th | 14th-29th | $p$ = .376 |
| Anxiety related to specific aspects of school (e.g. specific classes) | 39th/40th | 14th-29th | $p$ = .376 |
| Length of school day | 43rd-45th | 30th-98th | $p$ = .569 |
| Attendance pressure | 68th-71st | 30th-98th | $p$ = .756 |
| Changes to routine or staffing | 55th-59th | 30th-98th | $p$ = .688 |
| Structure of the school day | 78th-83rd | 30th-98th | $p$ = .858 |
| Lack of structure to the school day | 78th-83rd | 30th-98th | $p$ = .858 |
| Transition (within the school day) | 48th | 30th-98th | $p$ = .611 |
| Transition (from one year group to another) | 72nd-76th | 30th-98th | $p$ = .800 |
| Transition (from one school to another) | 49th-52nd | 30th-98th | $p$ = .634 |
| Covid-19 related changes to school day/environment | 78th-83rd | 30th-98th | $p$ = .858 |
| Journey to school e.g., school bus | 68th-71st | 30th-98th | $p$ = .756 |
| Excessive homework | 78th-83rd | 30th-98th | $p$ = .858 |
| Physical demands of school | 29th/30th | 14th-29th | $p$ = .689 |
| Problems with school regulations | 84th-98th | 30th-98th | - |
| An increasingly standardised education system | 19th | 14th-29th | $p$ = .992 |
| **Academic:** | | | |
| Academic pressure | 23rd/24th | 30th-98th | $p$ = .352 |
| Subject specific difficulties | 55th-59th | 30th-98th | $p$ = .688 |

| | | | |
|---|---|---|---|
| Difficulty accessing the curriculum | 31st | 30th-98th | *p* = .417 |
| **Disability-Related Factors:** | | | |
| Special Educational Needs (SEN) | 7th | 14th-29th | *p* = .433 |
| Ineffective SEN support | 5th | 30th-98th | *p* = .065 |
| Attention and learning difficulties | 25th | 30th-98th | *p* = .362 |
| Neurodivergence | 3rd | 14th-29th | *p* = .206 |
| Sensory processing difficulties/Sensory sensitivity | 6th | 6th-13th | *p* = .819 |
| Undiagnosed SEN/neurodivergence | 16th/17th | 6th-13th | *p* = .472 |
| Demand avoidance/heightened anxiety engaging in learning and/or adult-directed tasks | 4th | 14th-29th | *p* = .211 |
| Difficulty with social interactions and communication/social demands of school | 8th/9th | 14th-29th | *p* = .455 |
| Exhaustion from masking neurodivergence | 2nd | 14th-29th | *p* = .105 |
| Burnout (relating to neurodivergence) | 14th | 30th-98th | *p* = .230 |
| Being punished for behaviour relating to SEN/neurodivergence | 27th/28th | 30th-98th | *p* = .393 |
| Pressure to reduce/stop repetitive self-stimulatory behaviours (i.e. stimming) | 72nd-76th | 30th-98th | *p* = .800 |
| Not feeling their difficulties are believed | 27th/28th | 30th-98th | *p* = .393 |
| School not accommodating individual needs | 15th | 30th-98th | *p* = .242 |
| Cognitive rigidity | 61st-67th | 30th-98th | *p* = .720 |
| **Peer Relations:** | | | |
| Difficulties with friends/peers | 21st/22nd | 30th-98th | *p* = .333 |
| Unpredictable behaviour of peers/classmates | 32nd-34th | 30th-98th | *p* = .429 |
| Lack of friendships | 32nd-34th | 3rd-5th | *p* = .004** |
| Bullying (in-person) | 36th | 30th-98th | *p* = .455 |
| Bullying (cyber) | 77th | 30th-98th | *p* = .800 |
| Peer violence | 78th-83rd | 30th-98th | *p* = .858 |
| Sexual harassment/violence | 72nd-76th | 30th-98th | *p* = .800 |
| Peer/gang intimidation and/or exploitation at school | 72nd-76th | 30th-98th | *p* = .800 |
| Gang violence at school | 84th-98th | 30th-98th | - |

| | | | |
|---|---|---|---|
| **Pupil Behaviour:** | | | |
| Non-compliance | 68th-71st | 6th-13th | $p < .001$*** |
| Conduct disorder | 84th-98th | 30th-98th | - |
| Repeated sanctions (related to behaviour difficulties) | 55th-59th | 30th-98th | $p = .688$ |
| **Teacher Related Factors:** | | | |
| Lack of trust in teachers | 23rd/24th | 30th-98th | $p = .352$ |
| Difficulties with staff | 41st/42nd | 30th-98th | $p = .532$ |
| Teacher behaviour, e.g., shouting | 20th | 30th-98th | $p = .324$ |
| Lack of teacher training in SEN/neurodiversity | 29th/30th | 30th-98th | $p = .405$ |
| Inappropriate teacher training in SEN/neurodiversity | 37th | 30th-98th | $p = .469$ |
| Lack of teacher understanding (e.g. about the child or their neurodivergent diagnosis) | 10th | 30th-98th | $p = .145$ |
| **Reward/Puninshment by School Staff:** | | | |
| Use of behaviour modification strategies | 53rd-54th | 30th-98th | $p = .660$ |
| Repeated sanctions (related to attendance difficulties) | 84th-98th | 30th-98th | - |
| Use of restraint at school | 46th/47th | 30th-98th | $p = .589$ |
| Use of seclusion at school (i.e. forced to spend time alone against their will) | 55th-59th | 30th-98th | $p = .688$ |
| **Parent/Family Related Factors:** | | | |
| Poor parenting/lack of discipline | 84th-98th | 6th-13th | $p < .001$*** |
| Overprotective parenting | 84th-98th | 6th-13th | $p < .001$*** |
| To gain attention from a parent / caregiver / other | 84th-98th | 30th-98th | - |
| Difficulties at home/within the family | 84th-98th | 6th-13th | $p < .001$*** |
| Parental mental health | 84th-98th | 3rd-5th | $p < .001$*** |
| Detachment with little interaction among family members | 84th-98th | 30th-98th | - |
| Isolation with little interaction outside the family unit | 84th-98th | 30th-98th | - |
| Over-dependency within the family | 84th-98th | 14th-29th | $p < .001$*** |
| Child neglect | 78th-83rd | 30th-98th | $p = .858$ |
| **Other:** | | | |

| | | | |
|---|---|---|---|
| Poverty | 84th-98th | 30th-98th | - |
| Pursuit of tangible rewards outside of school | 84th-98th | 30th-98th | - |
| Preference for doing activities outside of school | 72nd-76th | 30th-98th | $p = .800$ |
| Athletic competition | 84th-98th | 30th-98th | - |
| Inability to engage in preferred routine | 68th-71st | 30th-98th | $p = .756$ |
| Other(s) | 49th-52nd | 14th-29th | $p = .122$ |
